# Metatranscriptomic Signatures of Lung Function in Pediatric Hematopoietic Cell Transplant Candidates

**DOI:** 10.1101/2021.09.21.21263910

**Authors:** Matt S. Zinter, Birgitta A. Versluys, Caroline A. Lindemans, Madeline Y. Mayday, Gustavo Reyes, Sara Sunshine, Marilynn Chan, Elizabeth Fiorino, Maria Cancio, Sabine Prevaes, Marina Sirota, Michael A. Matthay, Sandhya Kharbanda, Christopher C. Dvorak, Jaap J. Boelens, Joseph L. DeRisi

**Affiliations:** University of California, San Francisco, School of Medicine, Department of Pediatrics, Division of Critical Care Medicine; University of California, San Francisco, School of Medicine, Department of Pediatrics, Division of Allergy, Immunology, and Bone Marrow Transplantation; University Medical Center Utrecht, Department of Pediatric Stem Cell Transplantation; Princess Maxima Center for Pediatric Oncology, Department of Hematopoietic Cell Transplantation, Utrecht, The Netherlands; Yale University, Department of Pathology, Graduate Program in Experimental Pathology, and Yale Stem Cell Center; University of California, San Francisco, School of Medicine, Department of Biochemistry & Biophysics; University of California, San Francisco, School of Medicine, Department of Pediatrics, Division of Pulmonology; Cornell University, WC Medical College, Department of Pediatrics, Division of Pulmonology; Cornell University, WC Medical College, Department of Pediatrics; Memorial Sloan Kettering Cancer Center, Department of Pediatric Stem Cell Transplantation and Cellular Therapies; Department of Pediatric Pulmonology, Wilhelmina Children’s Hospital, University Medical Centre Utrecht, Utrecht University, Utrecht, The Netherlands; University of California, San Francisco, Bakar Computational Health Sciences Institute; University of California, San Francisco, School of Medicine, Department of Pediatrics; University of California, San Francisco, School of Medicine, Cardiovascular Research Institute, Departments of Medicine and Anesthesiology; Chan Zuckerberg Biohub, CA USA

**Keywords:** Respiratory Function Tests, Hematopoietic Stem Cell Transplantation, Microbiota, Bronchoalveolar Lavage, Pediatrics

## Abstract

**Rationale:** Impaired baseline lung function is associated with mortality after allogeneic hematopoietic cell transplantation (HCT). Limited knowledge of the molecular pathways that characterize pre-transplant lung function has hindered the development of lung-targeted interventions.

**Objectives:** To elucidate the biologic and microbiologic correlates of impaired lung function in pediatric allogeneic HCT candidates.

**Methods:** Between 2005-2016, 104 patients with malignant and non-malignant disorders ages 4-19 years underwent paired pulmonary function testing (PFT) and bronchoalveolar lavage (BAL) a median of 1-2 weeks prior to allogeneic transplant in Utrecht, the Netherlands. Cryopreserved BAL underwent RNA sequencing followed by alignment to microbial and human reference genomes for microbiome and gene expression analyses.

**Measurements and Main Results:** Abnormal pulmonary function was recorded in more than half the cohort, consisted most commonly of restriction and impaired diffusion, and was associated with both all-cause and lung-injury related mortality after HCT. BAL microbiome depletion of commensal supraglottic taxa such as *Haemophilus* and enrichment of nasal and skin taxa such as *Staphylococcus* were associated with worse measures of lung capacity and gas diffusion. In addition, impaired lung capacity and diffusion were also associated with gene expression signatures of alveolar epithelial proliferation, epithelial-mesenchymal transition, and downregulated immunity, suggesting a post-injury pro-fibrotic response. Detection of microbial depletion and abnormal epithelial gene expression in BAL enhanced the prognostic utility of pre-HCT PFTs for the outcome of post-HCT mortality.

**Conclusions:** These findings suggest a novel and potentially actionable connection between microbiome depletion, alveolar injury, and pulmonary fibrosis in the pathogenesis of pre-HCT lung dysfunction.

## INTRODUCTION

Allogeneic hematopoietic cell transplantation (HCT) is a life-saving cellular therapy used to cure underlying hematopoietic defects such as hematologic malignancies, primary immunodeficiencies, bone marrow failure syndromes, and hemoglobinopathies. While more than 7,500 children receive this treatment annually in the United States and Europe, the success of this therapy is limited by a broad range of post-HCT pulmonary injuries that develop in 10-40% of pediatric recipients in response to chemotherapeutic toxicity, infection, and impaired or dysregulated immunity (1–3). Comprehensive assessment of pulmonary health prior to HCT conditioning is critical in order to identify high-risk patients who may benefit from modified conditioning chemotherapy regimens, closer surveillance, and risk-stratified treatments aimed at mitigating the development of irreversible pulmonary toxicity (4–6).

In children, evaluation of pulmonary health prior to HCT is not standardized but may include symptom screening and physical exam, nasopharyngeal swab for common respiratory viruses, pulmonary function testing (PFT) if developmentally able (typically age ≥6 years), and targeted high-resolution chest tomography (HR-CT) and/or bronchoalveolar lavage (BAL) in patients with identified or suspected abnormalities (7, 8). As a sensitive metric of organ function, abnormal PFTs are detected in up to 60% of pediatric HCT candidates predominantly due to restriction and impaired diffusion (9–11) and are associated with the development of post-HCT complications including interstitial pneumonitis, pulmonary graft-versus-host disease (GVHD), and transplant-related mortality (TRM) (2, 3, 12–16).

To improve outcomes, it remains crucial to identify risk-factors for pre-HCT pulmonary dysfunction and elucidate pathobiologic correlates. To date, known risk-factors include exposure to alkylating and other pulmotoxic chemotherapy (e.g. busulfan, bleomycin, cyclophosphamide, carmustine, fludarabine, lomustine), thoracic radiation (particularly in concert with radiomimetics), thoracic surgery, chronic endothelial injury due to hemolysis and/or transfusion dependence, chronic inflammation, and recurrent pulmonary infections (17–19). Most patients with significantly impaired pre-HCT pulmonary function experience a number of these risk-factors, suggesting a multimodal pathogenesis. Despite the need for deeper understanding of the pathogenesis of pre-HCT pulmonary dysfunction, translational studies are limited by scarcity of investigable pulmonary samples from children.

In this study, we aimed to evaluate associations between pre-HCT pulmonary function and measures of pulmonary biology as assessed by metatranscriptomic RNA sequencing of paired BALs. We recently analyzed pre-HCT BAL samples from a cohort of pediatric HCT candidates and found that pre-HCT pulmonary bacterial depletion, viral enrichment, inflammation, and epithelial activation were associated with the development of post-HCT pulmonary toxicity (20). A subset of these patients were developmentally able to perform contemporaneous PFTs, presenting an opportunity to link the domains of organ function, biology, and microbiology within a cohort of high-risk patients. We hypothesized that the combination of pulmonary microbiome data paired with gene expression data could be associated with abnormal PFTs prior to HCT, thus providing further evidence for the pathobiologic role of the pulmonary microenvironment in pediatric HCT candidates.

## METHODS

### Study Design and Patients

As previously reported, pediatric allogeneic HCT candidates ≤19 years of age with malignant and non-malignant diseases underwent protocolized evaluation of pre-HCT pulmonary health between 2005-2016 in Utrecht, the Netherlands (11). This study included the subset who completed both pre-HCT PFTs and pre-HCT BAL prior to the start of conditioning chemotherapy. Informed consent was obtained with Institutional Review Board approval (#05/143 and #11/063). Details of approach to peri-HCT clinical care are described in **Supplemental Methods 1**.

### PFT Measurements

PFTs included spirometry, whole body plethysmography, and carbon monoxide diffusion capacity and were performed in developmentally capable children ages 4 and older according to established criteria (21, 22). Pre-bronchodilator PFT values of adequate performance quality were normalized for age, height, sex, and race and expressed as (1) a percentage deviation from the mean (% predicted) according to healthy Dutch children and (2) a standardized deviation from the mean (z-score) according to the Global Lung Index (GLI) (23, 24). Restriction, obstruction, mixed restriction/obstruction, diffusion impairment, and air trapping were identified first using conventional PFT cutoffs and were compared to GLI-recommended cutoffs wherein PFTs below the 5^th^ percentile (z-score <-1.64) were considered below the lower limit of normal (**Supplemental Methods 2**) (21, 24).

### BAL Measurements

BAL was performed in the absence of lower respiratory tract symptoms and at the time of another clinically-indicated procedure (e.g. bone marrow aspirate, central line placement) (19, 25). Cryopreserved aliquots of BAL underwent metatranscriptomic RNA sequencing as previously described (20). Briefly, samples underwent mechanical homogenization followed by RNA extraction and purification (26, 27). Sample RNA was combined with control spike-in RNA and underwent reverse transcription, library preparation, and 125nt paired-end sequencing on a NovaSeq 6000 instrument to a target depth of 40 million read-pairs per sample (**Supplemental Methods 3**). Resultant .fastq files underwent quality filtration followed by parallel alignment to both human and microbial reference genomes using the IDseq pipeline (28). Microbe count data were adjusted for laboratory contamination and estimated microbe mass was calculated using the reference spike-in (**Supplemental Methods 4**) (29).

### Data Analysis

Associations between PFTs and patient characteristics were tested using Kruskal-Wallis tests and Spearman correlations for categorical and continuous traits, respectively. Associations between pre-HCT PFTs and post-HCT outcomes were tested with Kaplan-Meier estimates and Cox proportional hazards models for all-cause mortality and cumulative incidence function with competing risks regression for fatal lung injury. The pulmonary microbiome was described using taxa correlation matrices and principal component analysis (PCA). Associations between PFTs and microbial masses were tested using negative binomial generalized linear models (NB-GLM) in the R statistical environment (edgeR) (30). Gene expression was characterized by calculating enrichment scores for the Hallmark Gene Sets in the Molecular Signatures Database (MSigDB) (31). Associations between PFTs and differentially expressed genes were tested using NB-GLM and characterized using pathway analysis (enrichR) (32). BAL cell type proportions and cell type-specific gene expression profiles were imputed using CiberSortX and the Human Lung Cell Atlas (**Supplemental Methods 5**) (33, 34).

### Data Sharing Statement

Sequencing files are available through dbGaP study accession phs002208.v1.p1.

## RESULTS

### Patient characteristics

Contemporaneous pre-HCT PFTs and BAL fluid were available for 104 patients. PFTs and BAL were typically performed 1-2 days apart and 1-2 weeks prior to HCT conditioning. Most patients were between 8-19 years of age, of self-reported Northern European ancestry, and preparing to undergo 1^st^ allogeneic HCT for underlying hematologic malignancy (**Table 1**). Most patients received myeloablative conditioning chemotherapy followed by an umbilical cord blood allograft. At 2-year post-HCT follow-up, 33 of 104 patients (31.7%) had died from relapsed disease or transplant toxicity.

**Table 1).**
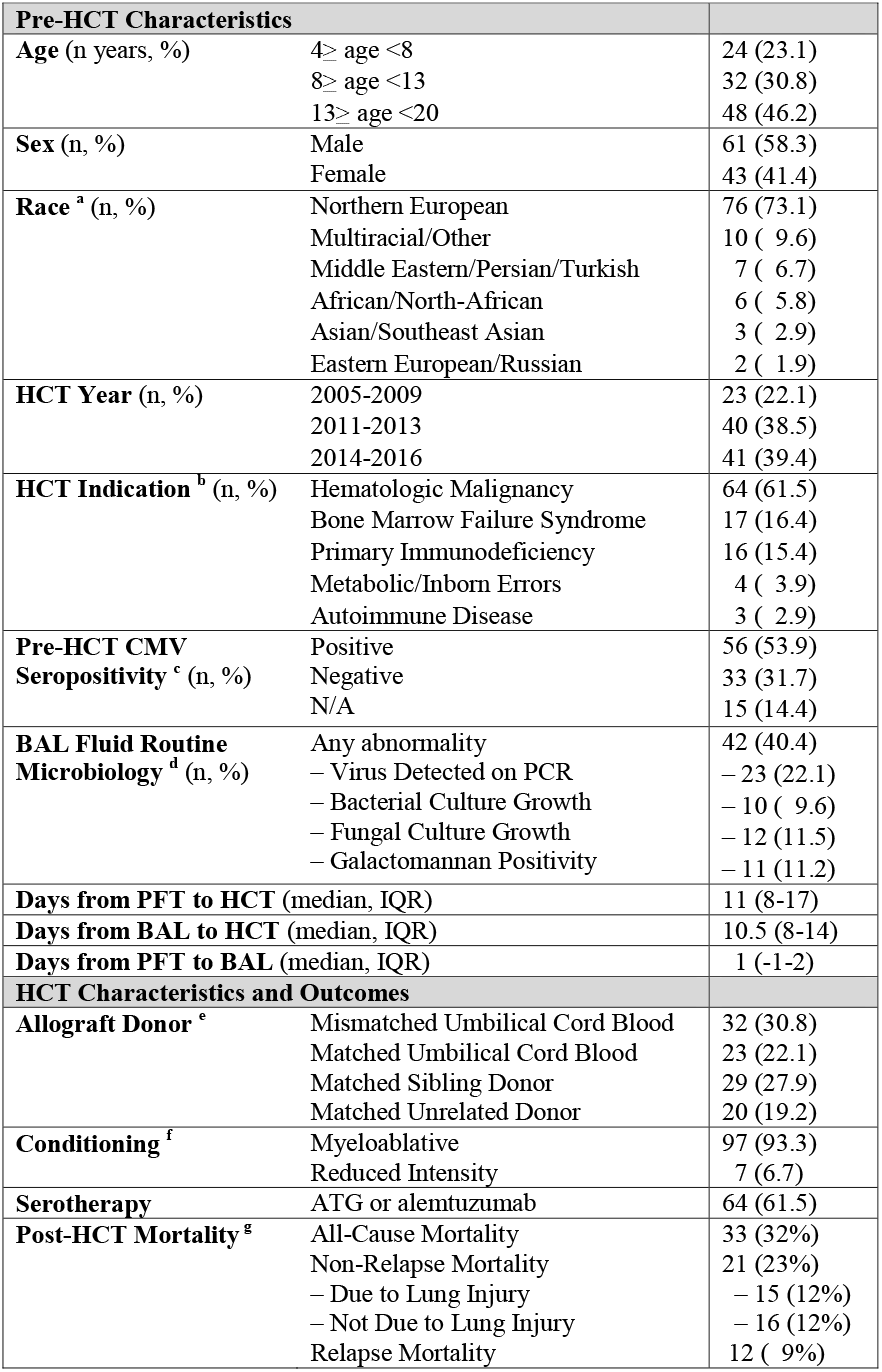
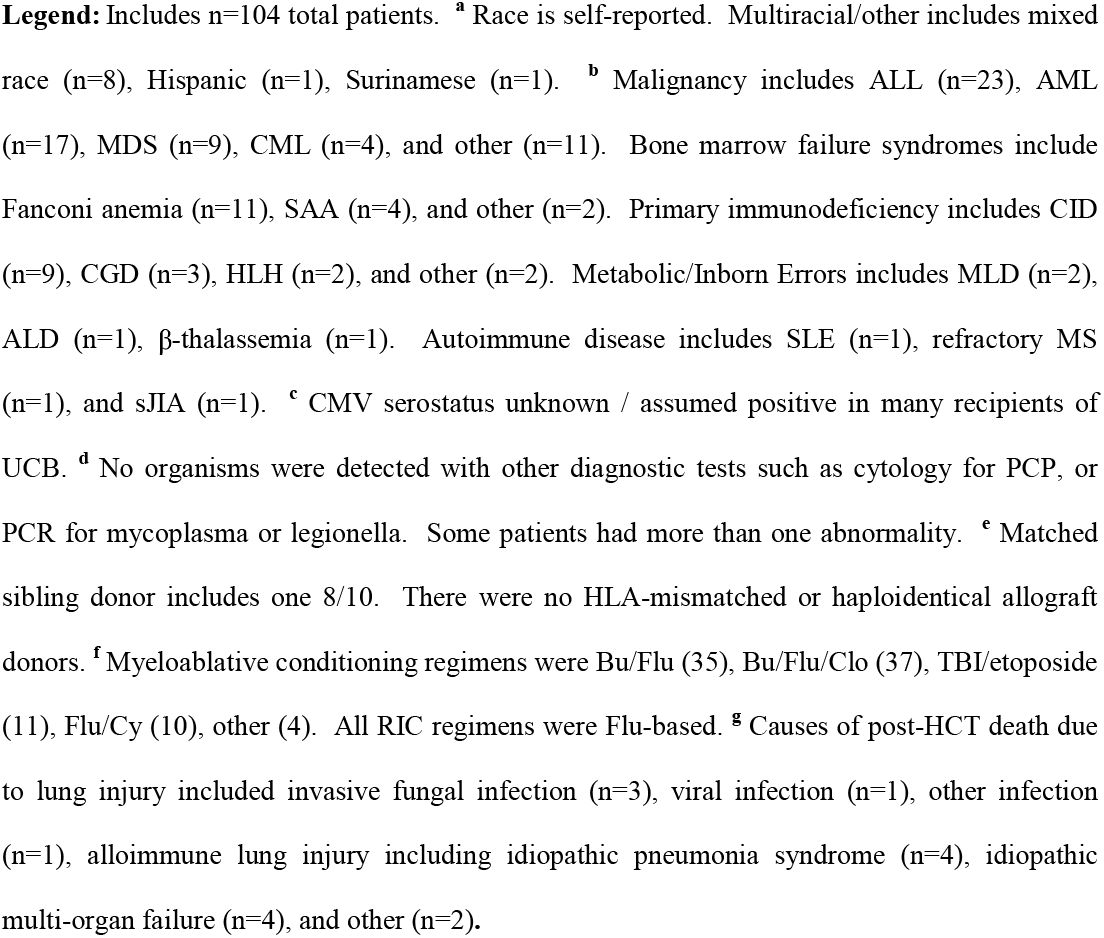
Pediatric Allogeneic HCT Candidate Patient Characteristics.

### Impaired Pulmonary Function Prior to HCT Is Common

All patients performed spirometry and a subset of typically older children performed plethysmography (n=78) and diffusion testing (n=63, **Table 2**). To determine the inter-relatedness of the different PFT measurements, we constructed a correlation matrix which showed broad grouping of PFTs into 4 clades pertaining to lung capacity, obstruction, diffusion, and air trapping (**Figure E1**). Using conventional cutoffs of absolute ratios and percentage deviations from the population mean (% predicted), PFT abnormalities were detected in 80 of 104 patients due to restriction (n=35/104), obstruction (n=12/104), mixed restriction/obstruction (n=1/104), impaired diffusion (n=21/63), and air trapping (n=44/78). Using GLI cutoffs based on standard deviations from the population mean (z-scores), PFT abnormalities were detected in 54 of 104 patients due to restriction (n=29/104), obstruction (n=7/104), impaired diffusion (n=14/63), and air trapping (n=16/78, **Table E1**). As GLI z-scores are not calculable for all measured PFTs, we used PFTs expressed as percentage of predicted for subsequent analyses.

**Table 2).** Pulmonary Function Tests in Pediatric Allogeneic HCT Candidates.

| **Spirometry <sup>a</sup> (n=104)** | **Actual** | **% of Predicted** | **Z-score** |
| --- | --- | --- | --- |
| Forced Vital Capacity (FVC, liters) | 2.21 (1.50-3.21) | 0.87 (0.74-0.96) | -1.07 (-2.28 to -0.35) |
| Forced Expiratory Volume in 1 second (FEV1, liters) | 1.94 (1.36-2.77) | 0.86 (0.75-0.96) | -1.14 (-2.08 to -0.40) |
| Forced Expiratory Volume in 1 second, adjusted (FEV1/FVC) | 0.88 (0.84-0.94) | 1.00 (0.95-1.06) | -0.17 (-0.83 to 0.92) |
| Peak Expiratory Flow (PEF, liters /second) | 4.44 (3.12-6.02) | 0.85 (0.76-0.97) | -- |
| Forced Expiratory Flow at 25% (FEF25, liters/second) | 3.85 (2.83-5.47) | 0.87 (0.72-0.99) | -- |
| Forced Expiratory Flow at 50% (FEF50, liters /second) | 2.58 (1.96-3.59) | 0.91 (0.69-1.05) | -- |
| Forced Expiratory Flow at 75% (FEF75, liters /second) | 1.23 (0.86-1.76) | 0.86 (0.62-1.06) | -0.40 (-1.28 to 0.32) |
| Forced Expiratory Flow 25%-75% (FEF25-75, liters/second) | 2.68 (1.73-3.81) | 0.98 (0.79-1.15) | -0.10 (-1.00 to 0.55) |
| **Plethysmography <sup>b</sup> (n=78)** |  |  |  |
| Total Lung Capacity (TLC, liters) | 3.75 (2.81-4.73) | 0.85 (0.74-0.95) | -0.76 (-1.45 to 0.09) |
| Vital Capacity (VC, liters) | 2.74 (1.94-3.33) | 0.90 (0.75-0.99) | -1.32 (-2.62 to -0.42) |
| Alveolar Volume (Va, liters) | 3.56 (2.69-4.63) | 0.91 (0.80-1.00) | -0.77 (-1.83 to -0.01) |
| Alveolar Volume/Total Lung Capacity (Va/TLC) | 0.95 (0.89-1.01) | 0.85 (0.93-0.97) | -- |
| Residual Volume (RV, liters) | 0.89 (0.71-1.35) | 0.99 (0.77-1.30) | 0.11 (-0.54 to 0.69) |
| Residual Volume/Total Lung Capacity (RV/TLC) | 0.26 (0.21-0.35) | 1.06 (0.83-1.44) | 0.51 (-0.11 to 1.47) |
| **Diffusion <sup>c</sup> (n=63)** |  |  |  |
| Diffusion Capacity for Carbon Monoxide (DLCO, mmol/min/kPa) | 5.78 (4.35-6.92) | 0.78 (0.68-0.95) | -1.65 (-2.53 to -0.34) |
| Diffusion Capacity for Carbon Monoxide/Va (DLCO/Va) | 1.50 (1.37-1.73) | 0.88 (0.76-1.02) | -0.69 (-1.58 to 0.12) |

**Legend:** <sup>a</sup> n=104 for spirometry, <sup>b</sup> n=78 for plethysmography, <sup>c</sup> n=63 for diffusion factor testing.
Median with interquartile ranges are listed. % of predicted based on *Koopman M et al* age- and sex-adjusted reference sets (23). Z-scores based on Global Lung Function Initiative reference values (24). All DLCO measurements are corrected for Hb.

### Abnormal Pulmonary Function Varies According to Age and Sex

We tested for clinical characteristics associated with PFT abnormalities and identified that older age was associated with restrictive lung disease as well as worse measures of obstruction (e.g. MEF_25_, MEF_50_, MEF_75_, and PEF, **Figure 1a**). Female sex was associated with worse measures of both lung capacity (e.g. FVC, Va) and diffusion (e.g. DLCO/Va) but better measures of obstruction (FEV_1_/FVC, **Figure 1b**). We tested for but did not identify an association between HCT indication and pre-HCT PFTs (**Supplemental Data**).

**Figure 1:**
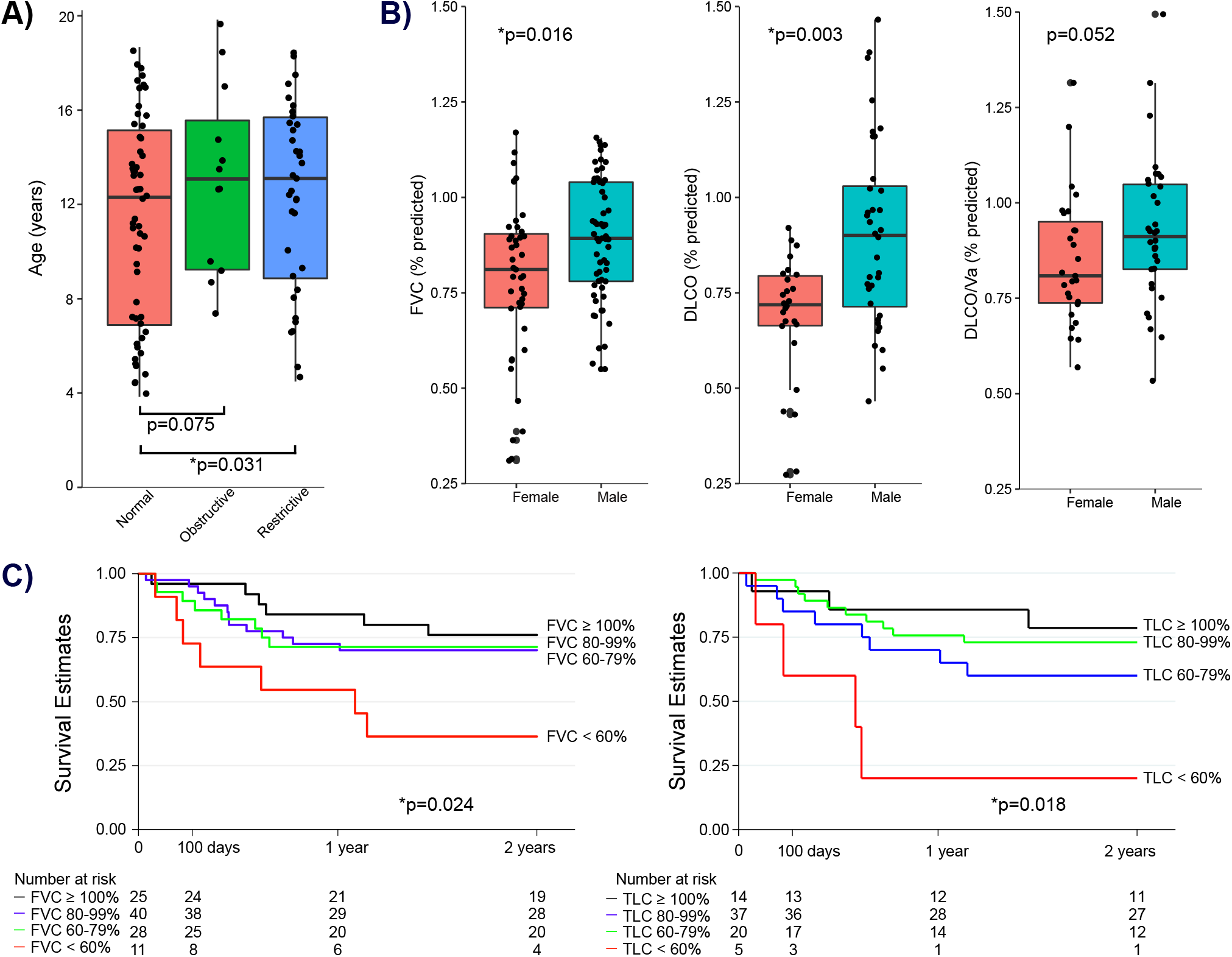
Association between Pre-HCT PFTs, Clinical Characteristics, and Post-HCT Outcomes. **(A)** Box-whisker plot of age distribution for patients with restriction, obstruction, or neither using conventional PFT categorization. Patients with either restriction or obstruction were older than patients with normal PFTs (Dunn’s Test p=0.031 and p=0.075, respectively). **(B)** Box-whisker plot of PFT distributions according to patient sex. Female patients had lower FVC_%pred_, DLCO_%pred_, and DLCO/Va_%pred_ (Wilcoxon rank-sum test p=0.016, p=0.003, p=0.052, respectively). **(C)** Kaplan-Meier post-HCT survival estimates according to pre-HCT FVC_%pred_ (left) and TLC_%pred_ (right). Both FVC_%pred_ and TLC_%pred_ were associated with post-HCT survival (Cox regression p=0.024 and p=0.018, respectively).

### Abnormal Pulmonary Function Is Associated with HCT Outcomes

Previous studies have shown impaired PFTs to be associated with post-HCT mortality (2, 3, 12–16). We confirmed these findings by demonstrating an association between reduced pre-HCT lung capacity and the development of all-cause mortality after HCT (HR 1.21 per 10% decline in FVC_%pred_, 95% CI 1.03-1.43, p=0.024, **Figure 1c**). To test whether these poor outcomes were due to acute lung injury specifically, we calculated cumulative incidence with competing risks regression and found that FVC_%pred_ was associated with the development of fatal lung injury in the first 180 days post-HCT (p<0.001, **Figure E2**).

### BAL Microbiome Composition Varies With Pulmonary Function

Our prior studies have associated alterations in the pre-HCT pulmonary microbiome composition with the later development of post-HCT outcomes (20). To determine whether the composition of the pre-HCT pulmonary microbiome is associated with lung function at the time of BAL measurement, we compared microbiome findings to contemporaneously performed PFTs. We first surveyed the composition of BAL microbiomes and found two oppositely correlated clusters of microbes: one containing common supraglottic taxa (e.g. *Haemophilus, Streptococcus, Neisseria*) and a second inversely correlated cluster containing common nasal, skin and environmental taxa (e.g. *Staphylococcus, Corynebacterium*, **Figure E3**). We represented these clusters mathematically using PCA and found that lower coordinates along principal component 1 (with top contributions from oropharyngeal taxa *Prevotella, Veillonella, Rothia*, and *Streptococcus*) were associated with worse measures of lung capacity (e.g. FVC, **Figure E4, Supplemental Data**).

Next, we used NB-GLM to identify specific taxa associated with each PFT. In models adjusted for patient age and sex, we found that lower BAL quantities of supraglottic taxa (e.g. *Haemophilus, Neisseria*) and greater BAL quantities of skin/environmental taxa (e.g. *Staphylococcus, Saccharomyces, Corynebacterium*) were associated with worse measures of lung capacity (FVC), obstruction (MEF25), diffusion impairment (DLCO), and air trapping (RV/TLC) (**Figure 2, Figure E5, Supplemental Data**). In addition, greater pulmonary fungal mass was associated with worse measures of lung capacity (e.g. FVC) and diffusion (e.g. DLCO), and greater mass of the Actinobacteria phylum, which includes the skin-associated genera *Corynebacterium, Micrococcus*, and *Cutibacterium*, was associated with worse measures of lung capacity (e.g. FVC) and obstruction (e.g. MEF25, MEF75). In contrast, lower mass of the Fusobacterium phylum was associated with worse measures of lung capacity (Va) and diffusion (DLCO). We did not find an association between PFTs and total viral RNA mass (**Supplemental Data**).

**Figure 2:**
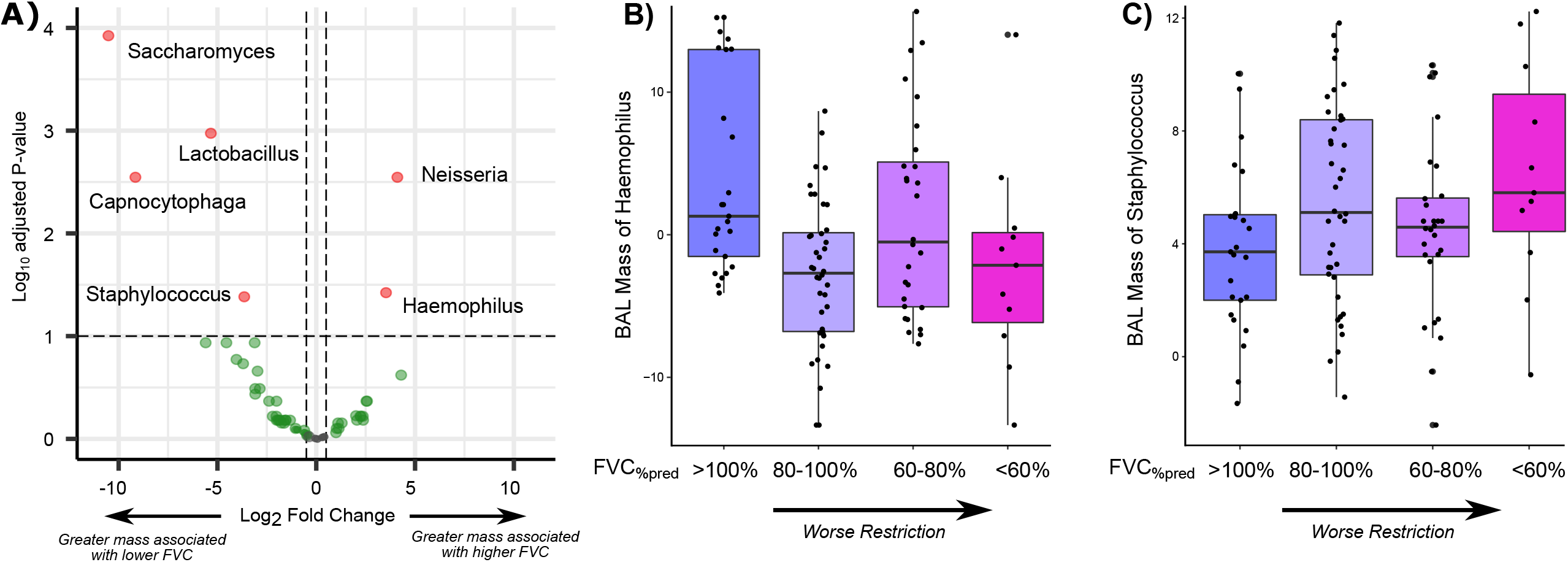
Association between BAL Microbiome and FVC_%pred_. **(A)** Volcano plot showing the association between BAL microbe mass and FVC_%pred_. Associations tested using negative binomial generalized linear models accounting for patient age and sex covariates. Positive log_2_ fold change (x-axis) indicates that greater microbial mass is associated with greater FVC_%pred_; negative log_2_ fold change indicates that greater microbial mass is associated with lower FVC_%pred_. **(B, C)** Box-whisker plot of mass of variance stabilizing transformed (*vst*) masses of Haemophilus (left) and Staphylococcus (right) according to category of FVC_%pred_. Worse restriction is associated with lower Haemophilus and greater Staphylococcus mass in BAL fluid (p-values calculated from NB-GLM as described above).

Some microbiome studies have suggested that aggregate microbiome traits such as alpha diversity, dominance, and richness, are associated with severity of diseases such as COPD and IPF (35, 36). In this cohort, neither alpha diversity nor richness were associated with PFTs. There was a trend towards an association between greater percentage of the microbiome occupied by the most abundance taxa (i.e. dominance) and lower measures of lung capacity (e.g. FVC, p=0.071).

### BAL Gene Expression Is Correlated with Pulmonary Function

To determine whether impaired pulmonary function was associated with unique gene expression signatures, we first utilized an overview approach and calculated BAL expression enrichment scores for each of the n=50 Hallmark Gene Sets in the Molecular Signatures Database. We found that measures of lung capacity and diffusion were positively associated with enrichment of pathways relating to immune activation, metabolism, and cell division and were negatively associated with pathways relating to epithelial mesenchymal transition and cell fate/differentiation (**Figure 3, Supplemental Data**).

**Figure 3:**
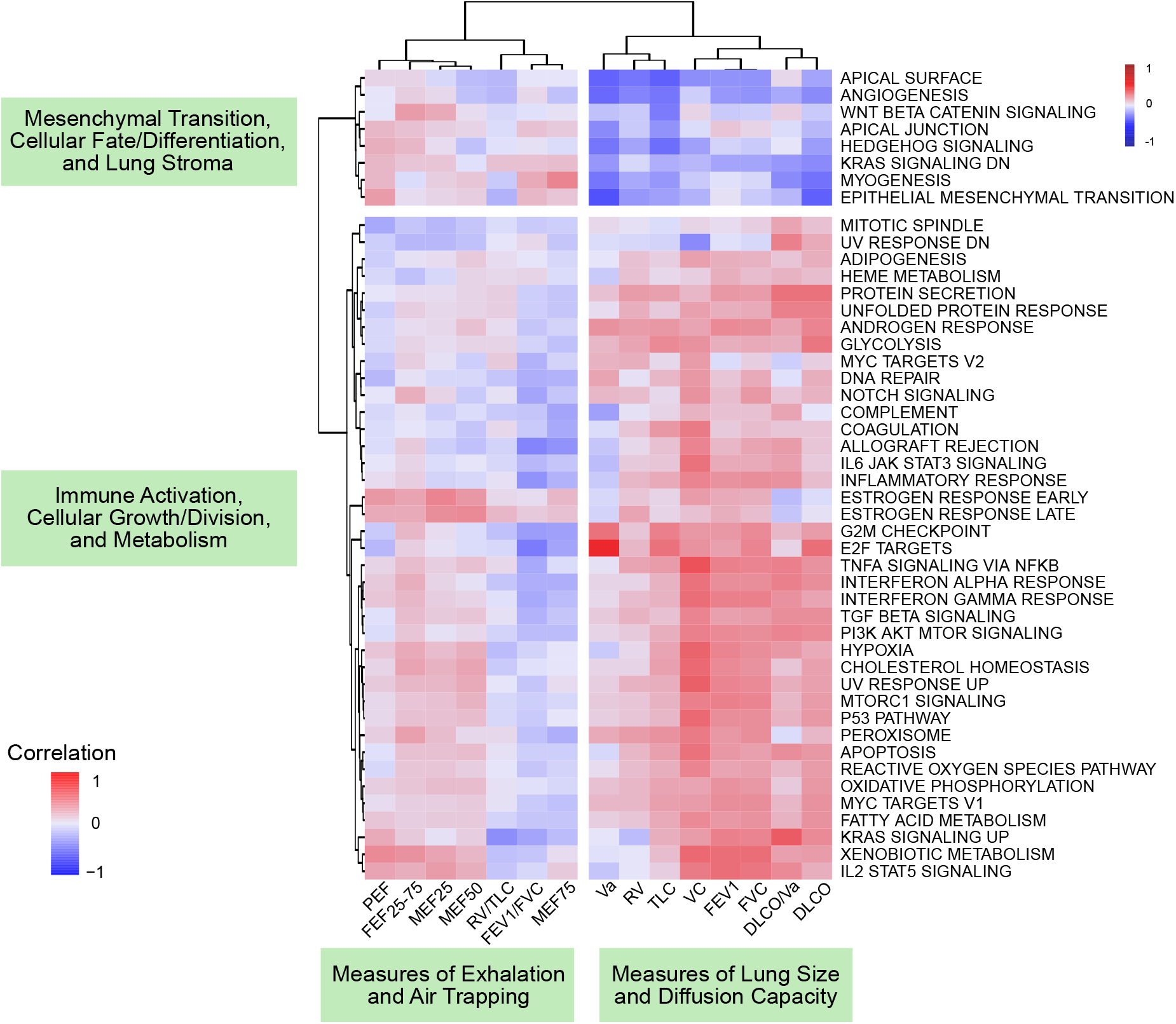
Association between BAL Hallmark Gene Expression and Pulmonary Function Testing. Hallmark gene set enrichment analysis using Poisson gene count distributions was used to calculate gene set enrichment scores in each patient. Spearman rank-based correlations between gene set enrichment scores (rows) and PFTs (columns) were calculated and plotted (cells) with hierarchical clustering of columns and rows according to Euclidean distance. Dendrogram-based clustering shows that PFT measures of lung size and diffusion were positively associated with a cluster of hallmark gene sets corresponding to immune activation and cellular growth, division, and metabolism and were negatively associated with a cluster of hallmark gene sets corresponding to mesenchymal transition, cellular fate/differentiation, and the lung stroma.

We next tested for specific genes whose expression levels were associated with each PFT. Pathway analysis showed that lower levels of both FVC and DLCO/Va were associated with greater expression of pulmonary epithelial genes, including surfactant genes (e.g. SFTPA1/2, SFTPC, SFTPD), keratinization genes (e.g. KRTAP3.2, KRTAP5.6), and epithelial sensory/stimulus response genes (e.g. OR2A1, OR4E2), as well as lower expression of innate immunity genes (e.g. CXCL8, HLA-DRA, IL1B, LYZ; **Figure 4a, 4b**). In contrast, lower FEV1/FVC was associated with greater expression of innate immunity genes and lower expression of alveolar epithelial genes. These findings persisted after adjusting models for age and sex. Few genes (n=9) were associated with a metric of air trapping (RV/TLC, **Supplemental Data**).

**Figure 4:**
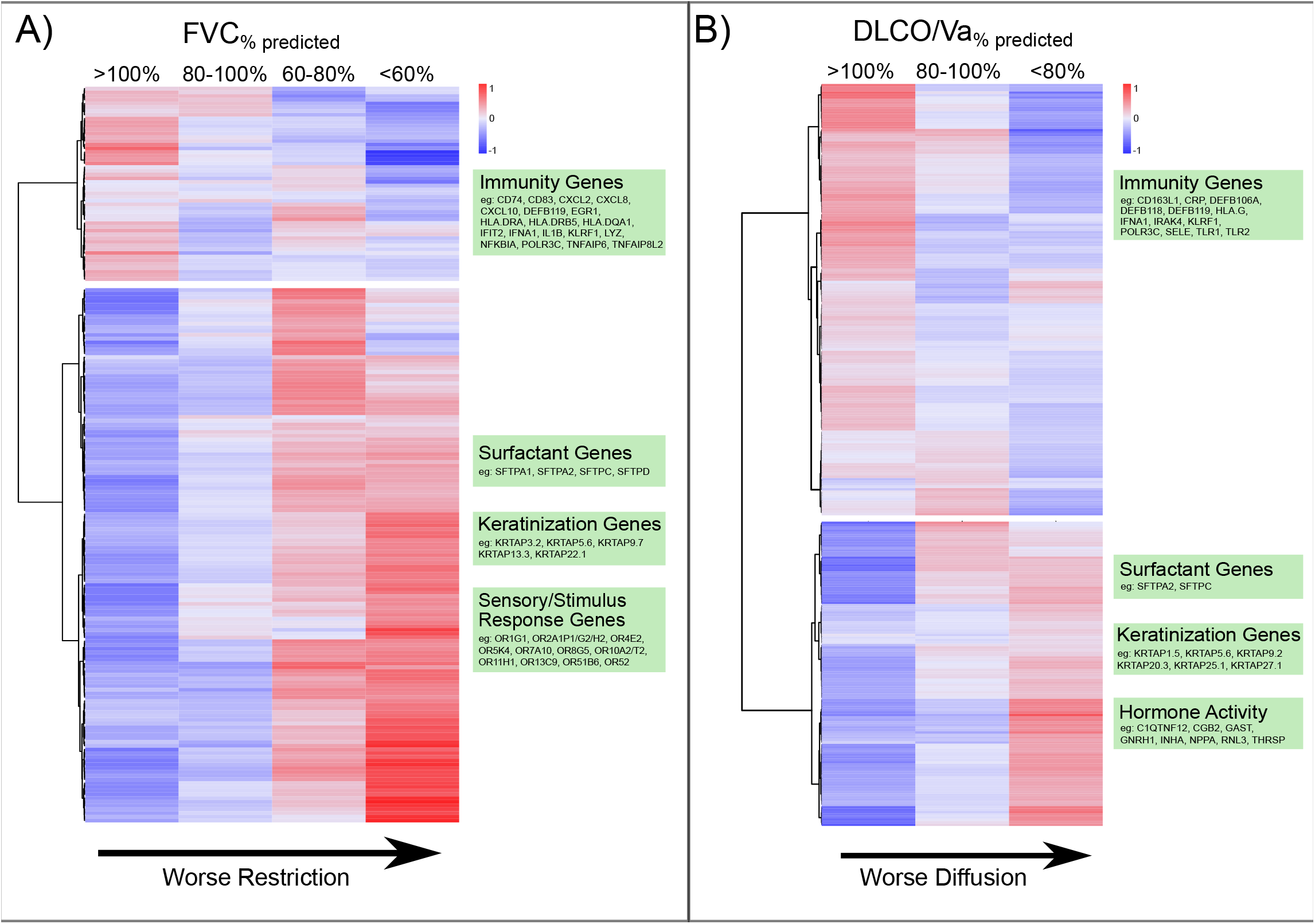
Association between BAL Gene Expression and Pulmonary Function Testing. Heatmaps of differential gene expression according to pulmonary restriction (**A**) and impaired diffusion (**B**). Negative binomial models were used to identify differentially expressed genes (rows) across the continuum of FVC_%pred_ and DLCO/Va_%pred_ (columns). Cell shading indicates mean gene expression for each PFT subgroup after variance stabilizing transformation, centering, and scaling. Dendrogram-based clustering shows that declining FVC_%pred_ was associated with a decrease in expression of immune-related genes and an increase in expression of airway epithelial genes relating to surfactant, keratinization, and olfactory response. Declining DLCO/Va_%pred_ was also associated with a decrease in expression of immune-related genes and an increase in expression of airway epithelial genes as well as hormone response genes.

### BAL Cell Fraction is Associated with Pulmonary Function

Since BAL samples are heterogeneous cell mixtures, we posited that these results could be due to either different proportions of cell types within samples, different expression patterns of each cell type within samples, or both. Therefore, we used *in silico* cell deconvolution techniques based on the Human Lung Cell Atlas to impute lung cell proportions in each sample. BAL cell types clustered into 3 groups consisting of innate immune cells, upper airway cells, and lower airway and adaptive immune cells (**Figure 5a**). Patients with restriction had greater BAL enrichment scores for type 1 and 2 alveolar epithelial cells (AECs) (**Figure 5b**). In addition, patients with impaired diffusion had greater BAL enrichment scores for type 2 AECs (**Supplemental Data)**. Next, using the CiberSortX algorithm (33), we estimated the average gene expression profile for each BAL cell type. In patients with restriction, type 1 AECs showed increased transcription of genes related to epithelial-mesenchymal transition, hedgehog signaling, and KRAS signaling, and T- and B-lymphocytes showed an overall decrease in transcription of genes relating to growth and inflammation (**Supplemental Data**). We did not detect differences in innate immune cell or upper airway epithelial cell-specific gene expression among patients with vs. without restriction.

**Figure 5:**
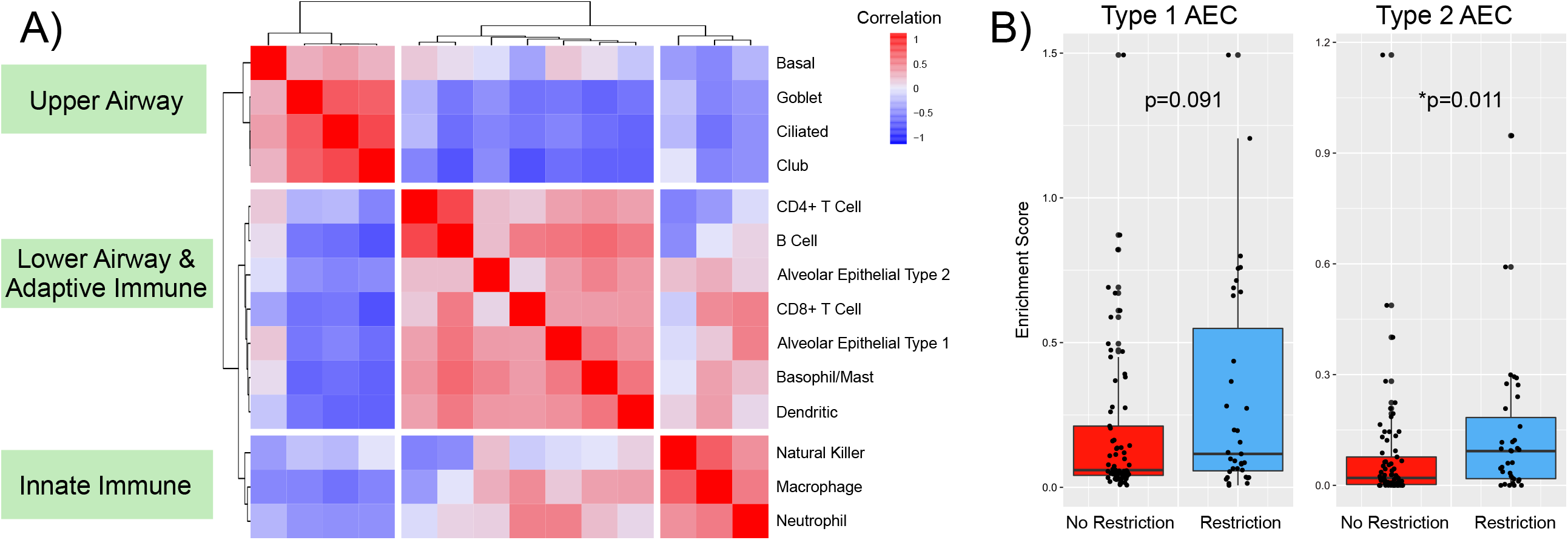
BAL Cell Type Differential. Single cell RNA sequencing data from The Human Lung Cell Atlas was used to calculate cell-type specific marker genes and impute cell-type fractions and enrichment scores (*CiberSortX*). **(A)** Highly correlated cell types were identified by calculating Spearman rank-based correlation of cell type fractions followed by hierarchical clustering of correlation distances. **(B)** Cell type enrichment scores were tested for association with pulmonary restriction using Kruskal-Wallis testing.

### Interactions Between PFTs, BAL Metatranscriptome, and Post-HCT Mortality

Given prior findings that pre-HCT BAL metatranscriptomes are associated with post-HCT mortality (20), we next sought to determine whether these associations are independent of measured pulmonary function. Using multivariable modeling, we found that lower BAL mass of oropharyngeal taxa (e.g. *Haemophilus, Neisseria*) was associated with post-HCT all-cause mortality independent of FVC, age, and sex ((**Supplemental Data**). We identified a statistical interaction whereby progressively lower BAL mass of oropharyngeal taxa strengthened the association between poor pre-HCT FVC and post-HCT mortality. In addition, lower BAL expression of genes related to airway epithelial cell integrity, polarity, and junction formation (e.g. Hallmark Apical Surface) were also associated with post-HCT mortality independent of FVC, age, and sex. The addition of BAL *Haemophilus* mass and BAL Hallmark Apical Surface gene set enrichment to a Cox survival model of FVC, age, and sex significantly improved model performance across strata of lung function (likelihood ratio test p=0.002, **Figure 6**).

**Figure 6:**
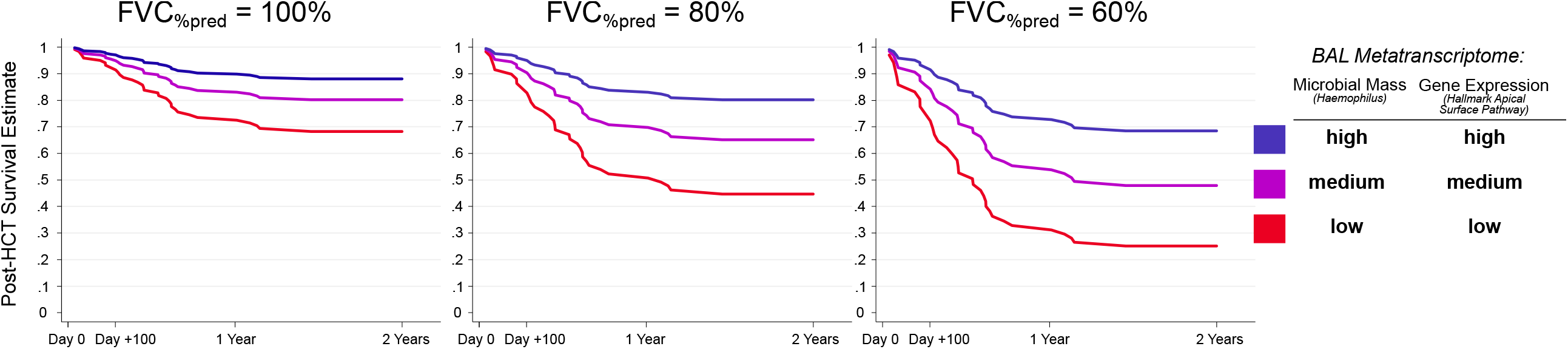
Survival Estimates According to Pre-HCT BAL Metatranscriptome and Pre-HCT FVC. A multivariable Cox regression model for the outcome of post-HCT all cause mortality was fit using pre-HCT FVC_%pred_, pre-HCT BAL mass of *Haemophilus* (*vst* transformed), and pre-HCT BAL enrichment score of MSigDB Hallmark Apical Surface Gene Set with covariate adjustment for patient age and sex. Mortality estimates are plotted according to pre-HCT FVC (100% vs 80% vs 60%) and high, medium, and low *Haemophilus* mass and Apical Surface gene expression. High, medium, and low refer to 75^th^, 50^th^, and 25^th^ percentile within the cohort.

## DISCUSSION

The primary finding of this study is that in pediatric HCT candidates, pulmonary microbiome and gene expression signatures are associated with pulmonary function and may augment the utility of PFTs in the stratification of post-HCT mortality risk. Specifically, we found that the pulmonary microbiomes of patients with restriction and diffusion impairment were depleted of common supraglottic taxa and instead enriched in skin, nasal, and environmental taxa. In addition, the pulmonary transcriptomes of patients with restriction and diffusion impairment showed increased activation of alveolar epithelial cells and a deficit in immune cell signaling consistent with post-injury fibrotic transformation. Further, BAL signatures of commensal microbial depletion and diminished epithelial homeostasis were associated with post-HCT mortality independent of pre-HCT pulmonary function. Together these findings suggest a novel connection between lung function, biology, and microbiology in pediatric HCT candidates and suggest possible biologic targets to optimize lung function in children preparing to undergo HCT.

First, our finding that pulmonary microbiome depletion is associated with impaired lung function in pediatric HCT candidates is novel and supports our previous finding that pre-HCT pulmonary microbiome depletion is associated with post-HCT lung injury (20). Although studies of the pulmonary microbiome in children are rare, investigations of the more readily-studied intestinal microbiome have similarly shown that loss of intestinal biodiversity and biomass are associated with organ dysfunction as well as poor post-HCT outcomes (37). Whether pulmonary microbial dysbiosis can directly contribute to lung disease or is simply a biomarker of other exposures remains unclear. On the one hand, microbial dysbiosis is linked to antimicrobial exposure, which is more common in patients with underlying diseases such as malignancy; such patients have confounding reasons for poor lung function such as greater exposure to pulmotoxic chemotherapy. However, emerging studies suggest that microbial dysbiosis disrupts the control of pathogen overgrowth and contributes to a loss of immune tolerance at the epithelial-parenchymal boundary, and therefore reciprocal causality between lung dysfunction and microbiome disturbances seems likely (35, 38). To that end, we found that pulmonary biomass of supraglottic commensals was inversely related to biomass of skin, nasal, and environmental taxa, supporting the theory that maintenance of the respiratory microbiomes may exclude invasion by outside organisms. Ultimately, strategies to modulate the lung microbiome in order to improve pulmonary health remain in development and include investigation of both ingested and inhaled probiotics and other microbial metabolites (39, 40).

A second finding of this work is the association between altered alveolar epithelial gene expression, restriction, and impaired diffusion. By combining differential gene expression with cell type imputation, we identified a greater proportion of type 1 and 2 AECs in patients with restriction and diffusion and cell-type specific gene expression suggested an increased transcriptional state of these cells, associated with growth factors and increased kinase expression. The increased expression of surfactant genes and overall AEC activity is consistent with response to alveolar injury and has been demonstrated in numerous cohorts of lung-injured patients (41, 42). By combining hallmark gene set enrichment with these findings, we found that pathways related to the lung stroma were significantly upregulated, including angiogenesis, myogenesis, epithelial mesenchymal transition, and cell fate (Wnt/β-catenin signaling and hedgehog signaling). Activation of stromal pathways in the lung has been associated with a pro-fibrotic signaling state and has been identified in cohorts of patients with IPF, thus providing a plausible mechanistic link between these gene expression findings and the observed impairment in PFTs (43, 44). As lung-resident mesenchymal stem cells undergoing myofibroblastic differentiation strongly express the Wnt/β-catenin and hedgehog signaling pathways, anti-fibrotic agents targeting this pathway such as pirfenidone remain of utmost interest (45, 46).

A third finding of this work is that patients with pulmonary restriction and impaired diffusion displayed an overall deficit in lower airway immune signaling. In addition to lower overall expression of immune genes among patents with restriction, cell type deconvolution inferred a less active transcriptional state for lymphocytes specifically. It seems counterintuitive that patients with poor PFTs would have a relative deficit in pulmonary immune activity, especially given our prior findings that pre-HCT pulmonary inflammation was associated with post-HCT lung injury. There are three potential explanations. First, the latter finding of pulmonary inflammation was tied to pre-HCT viral infection, which was more prevalent in children <4 years old. As younger children are not typically able to perform PFTs, they were excluded from this present study. Second, while pulmonary inflammation is a hallmark of acute lung injury including chemoradiation-induced lung injury, it typically subsides after the initial insult and is followed by a profound anti-inflammatory/pro-fibrotic phase (47), which more likely represents the phase of injury for the patients in this study. Third, patients with impaired lower airway immunity may be at greater risk for infections and associated sequelae such as post-infectious fibrosis. Ultimately, the significance of impaired pulmonary immune environment in pediatric HCT candidates with poor lung function needs to be explored with novel approaches in the future.

A fourth and unanticipated finding of this work was the identification of significant sex differences in pulmonary function. Although both male and female study participants had worse lung function than age- and sex-matched controls, females had profoundly worse measures of both restriction and impaired diffusion and males had slightly worse measures of obstruction. Long-term follow-up studies of pediatric oncology and HCT patients have also demonstrated associations between female sex, restriction, and impaired diffusion as well as between male sex and obstruction; this study adds to the field by identifying these disparities prior to HCT. A likely culprit for these toxicities are underlying differences in chemotherapy pharmacokinetics and pharmacodynamics, which merit exploration in future studies (48–50).

Our study has several strengths. First, the analysis of contemporaneously measured PFTs and BAL is unique among pediatric cohorts and allows novel insights into lung function, biology, and microbiology. Second, PFT measurements included GLI Z-scores according to updated ATS/ERS recommendations. Third, we analyzed BAL using a paired assessment of microbiome and gene expression. Several limitation merit comment. First, as with all observational human studies, we are unable to prove mechanism or causation among the factors measured and described herein. Second, we were unable to ascertain all possible risk-factors that might have influenced the pulmonary microenvironment, including history of pulmonary infections, prior antimicrobial exposure, and detailed chemotherapeutic regimens. Finally, younger children were necessarily excluded due to inability to perform PFTs. However, given our findings associating PFTs with lower respiratory biology, this study raises the possibility that BAL RNA sequencing could be a useful surrogate to detect pro-fibrotic states in those unable to perform PFTs.

In conclusion, in this study of pediatric HCT candidates, predominant patterns of pulmonary restriction and impaired gas diffusion were associated with microbiome depletion, proliferation of the alveolar epithelium, and a deficit in immune signaling. Both pre-HCT PFTs and BAL metatranscriptomes were independently associated with post-HCT mortality. These findings significantly increase our knowledge of the pathobiology of lung dysfunction in patients preparing to undergo HCT and offer numerous potential biologic targets that might be manipulated to optimize lung health in pediatric HCT patients.

## Data Availability

Data Sharing Statement: Sequencing files are available through dbGaP study accession phs002208.v1.p1.

## ACKNOWLEDGEMENTS

We thank Dr. Dean Sheppard, MD for his critical review of this manuscript.

## Notes

### Competing Interest Statement

The authors have declared no competing interest.

### Funding Statement

NHLBI K23HL146936, NICHD K12HD000850, American Thoracic Society Foundation Grant

### Author Declarations

University Medical Center Utrecht IRB #05/143 and #11/063

## REFERENCES

1. D’Souza A, Fretham C, Lee SJ, Aurora M, Brunner J, Chhabra S, Devine S, Eapen M, Hamadani M, Hari P, Pasquini MC, Phelan RA, Riches ML, Rizzo JD, Saber W, Shaw BE, Spellman SR, Steinert P, Weisdorf DJ, Horowitz MM. Current Use and Trends in Hematopoietic Cell Transplantation in the United States. BiolBlood Marrow Transplant 2020;26:e177–182.

2. Broglie L, Fretham C, Al-Seraihy A, George B, Kurtzberg J, Loren A, MacMillan M, Martinez C, Davies SM, Pasquini MC. Pulmonary Complications in Pediatric and Adolescent Patients Following Allogeneic Hematopoietic Cell Transplantation. Biol Blood Marrow Transplant 2019;25:2024–2030.

3. Kaya Z, Weiner DJ, Yilmaz D, Rowan J, Goyal RK. Lung function, pulmonary complications, and mortality after allogeneic blood and marrow transplantation in children. Biol Blood Marrow Transplant 2009;15:817–826.

4. Tamburro RF, Cooke KR, Davies SM, Goldfarb S, Hagood JS, Srinivasan A, Steiner ME, Stokes D, DiFronzo N, El-Kassar N, Shelburne N, Natarajan A, Pulmonary Complications of Pediatric Hematopoietic Stem Cell Transplantation Workshop Participants. Pulmonary Complications of Pediatric Hematopoietic Stem Cell Transplantation (HCT): An NIH Workshop Summary. Ann Am Thorac Soc 2020;doi:10.1513/AnnalsATS.202001-006OT.

5. Duque-Afonso J, Ihorst G, Waterhouse M, Zeiser R, Wäsch R, Bertz H, Yücel M, Köhler T, Müller-Quernheim J, Marks R, Finke J. Comparison of reduced-toxicity conditioning protocols using fludarabine, melphalan combined with thiotepa or carmustine in allogeneic hematopoietic cell transplantation. Bone Marrow Transplant 2021;56:110–120.

6. Schechter T, Naqvi A, Weitzman S. Risk for complications in patients with hemophagocytic lymphohistiocytosis who undergo hematopoietic stem cell transplantation: myeloablative versus reduced-intensity conditioning regimens. Expert Rev Clin Immunol 2014;10:1101–1106.

7. Cheng G-S. Pulmonary Function and Pretransplant Evaluation of the Hematopoietic Cell Transplant Candidate. Clinics in Chest Medicine 2017;38:307–316.

8. Chien JW, Madtes DK, Clark JG. Pulmonary function testing prior to hematopoietic stem cell transplantation. Bone Marrow Transplant 2005;35:429–435.

9. Wieringa J, van Kralingen KW, Sont JK, Bresters D. Pulmonary function impairment in children following hematopoietic stem cell transplantation. Pediatr Blood Cancer 2005;45:318–323.

10. Smith AR, Majhail NS, MacMillan ML, DeFor TE, Jodele S, Lehmann LE, Krance R, Davies SM. Hematopoietic cell transplantation comorbidity index predicts transplantation outcomes in pediatric patients. Blood 2011;117:2728–2734.

11. Versluys AB, van der Ent K, Boelens JJ, Wolfs T, de Jong P, Bierings MB. High Diagnostic Yield of Dedicated Pulmonary Screening before Hematopoietic Cell Transplantation in Children. Biol Blood Marrow Transplant 2015;21:1622–1626.

12. Srinivasan A, Srinivasan S, Sunthankar S, Sunkara A, Kang G, Stokes DC, Leung W. Pre-hematopoietic stem cell transplant lung function and pulmonary complications in children. Ann Am Thorac Soc 2014;11:1576–1585.

13. Ginsberg JP, Aplenc R, McDonough J, Bethel J, Doyle J, Weiner DJ. Pre-transplant lung function is predictive of survival following pediatric bone marrow transplantation. Pediatr Blood Cancer 2010;54:454–460.

14. Quigg TC, Kim Y-J, Goebel WS, Haut PR. Lung function before and after pediatric allogeneic hematopoietic stem cell transplantation: a predictive role for DLCOa/VA. J Pediatr Hematol Oncol 2012;34:304–309.

15. Herasevich S, Frank RD, Bo H, Alkhateeb H, Hogan WJ, Gajic O, Yadav H. Pretransplant Risk Factors Can Predict Development of Acute Respiratory Distress Syndrome after Hematopoietic Stem Cell Transplantation. Ann Am Thorac Soc 2021;18:1004–1012.

16. Lee HJ, Kim K, Kim SK, Lee JW, Yoon J-S, Chung N-G, Bin C. Hb-adjusted DLCO with GLI reference predicts long-term survival after HSCT in children. Bone Marrow Transplant 2021;56:1929–1936.

17. Huang T-T, Hudson MM, Stokes DC, Krasin MJ, Spunt SL, Ness KK. Pulmonary outcomes in survivors of childhood cancer: a systematic review. Chest 2011;140:881–901.

18. Kharbanda S, Panoskaltsis-Mortari A, Haddad IY, Blazar BR, Orchard PJ, Cornfield DN, Grewal SS, Peters C, Regelmann WE, Milla CE, Baker KS. Inflammatory cytokines and the development of pulmonary complications after allogeneic hematopoietic cell transplantation in patients with inherited metabolic storage disorders. Biol Blood Marrow Transplant 2006;12:430–437.

19. Versluys B, Bierings M, Murk JL, Wolfs T, Lindemans C, Vd Ent K, Boelens JJ. Infection with a respiratory virus before hematopoietic cell transplantation is associated with alloimmune-mediated lung syndromes. J Allergy Clin Immunol 2018;141:697–703.

20. Zinter MS, Lindemans CA, Versluys BA, Mayday MY, Sunshine S, Reyes G, Sirota M, Sapru A, Matthay MA, Kharbanda S, Dvorak CC, Boelens JJ, DeRisi JL. The pulmonary metatranscriptome prior to pediatric HCT identifies post-HCT lung injury. Blood 2021;137:1679–1689.

21. Stanojevic S, Graham BL, Cooper BG, Thompson BR, Carter KW, Francis RW, Hall GL, Global Lung Function Initiative TLCO working group, Global Lung Function Initiative (GLI) TLCO. Official ERS technical standards: Global Lung Function Initiative reference values for the carbon monoxide transfer factor for Caucasians. Eur Respir J 2017;50:.

22. Culver BH, Graham BL, Coates AL, Wanger J, Berry CE, Clarke PK, Hallstrand TS, Hankinson JL, Kaminsky DA, MacIntyre NR, McCormack MC, Rosenfeld M, Stanojevic S, Weiner DJ, ATS Committee on Proficiency Standards for Pulmonary Function Laboratories. Recommendations for a Standardized Pulmonary Function Report. An Official American Thoracic Society Technical Statement. Am J Respir Crit Care Med 2017;196:1463–1472.

23. Koopman M, Zanen P, Kruitwagen CLJJ, van der Ent CK, Arets HGM. Reference values for paediatric pulmonary function testing: The Utrecht dataset. Respir Med 2011;105:15–23.

24. Quanjer PH, Stanojevic S, Cole TJ, Baur X, Hall GL, Culver BH, Enright PL, Hankinson JL, Ip MSM, Zheng J, Stocks J, ERS Global Lung Function Initiative. Multi-ethnic reference values for spirometry for the 3-95-yr age range: the global lung function 2012 equations. Eur Respir J 2012;40:1324–1343.

25. Versluys AB, Rossen JW, van Ewijk B, Schuurman R, Bierings MB, Boelens JJ. Strong association between respiratory viral infection early after hematopoietic stem cell transplantation and the development of life-threatening acute and chronic alloimmune lung syndromes. Biol Blood Marrow Transplant 2010;16:782–791.

26. Zinter MS, Dvorak CC, Mayday MY, Iwanaga K, Ly NP, McGarry ME, Church GD, Faricy LE, Rowan CM, Hume JR, Steiner ME, Crawford ED, Langelier C, Kalantar K, Chow ED, Miller S, Shimano K, Melton A, Yanik GA, Sapru A, DeRisi JL. Pulmonary Metagenomic Sequencing Suggests Missed Infections in Immunocompromised Children. Clin Infect Dis 2019;68:1847–1855.

27. Mayday MY, Khan LM, Chow ED, Zinter MS, DeRisi JL. Miniaturization and optimization of 384-well compatible RNA sequencing library preparation. PLoS One 2019;14:e0206194.

28. Kalantar KL, Carvalho T, de Bourcy CFA, Dimitrov B, Dingle G, Egger R, Han J, Holmes OB, Juan Y-F, King R, Kislyuk A, Lin MF, Mariano M, Morse T, Reynoso LV, Cruz DR, Sheu J, Tang J, Wang J, Zhang MA, Zhong E, Ahyong V, Lay S, Chea S, Bohl JA, Manning JE, Tato CM, DeRisi JL. IDseq-An open source cloud-based pipeline and analysis service for metagenomic pathogen detection and monitoring. Gigascience 2020;9:giaa111.

29. Zinter MS, Mayday MY, Ryckman KK, Jelliffe-Pawlowski LL, DeRisi JL. Towards precision quantification of contamination in metagenomic sequencing experiments. Microbiome 2019;7:62–6.

30. Anders S, McCarthy DJ, Chen Y, Okoniewski M, Smyth GK, Huber W, Robinson MD. Count-based differential expression analysis of RNA sequencing data using R and Bioconductor. NatProtoc 2013;8:1765–1786.

31. Liberzon A, Birger C, Thorvaldsdóttir H, Ghandi M, Mesirov JP, Tamayo P. The Molecular Signatures Database (MSigDB) hallmark gene set collection. Cell Syst 2015;1:417–425.

32. Chen EY, Tan CM, Kou Y, Duan Q, Wang Z, Meirelles GV, Clark NR, Ma’ayan A. Enrichr: interactive and collaborative HTML5 gene list enrichment analysis tool. BMC Bioinformatics 2013;14:128–128.

33. Newman AM, Steen CB, Liu CL, Gentles AJ, Chaudhuri AA, Scherer F, Khodadoust MS, Esfahani MS, Luca BA, Steiner D, Diehn M, Alizadeh AA. Determining cell type abundance and expression from bulk tissues with digital cytometry. Nat Biotechnol 2019;37:773–782.

34. Travaglini KJ, Nabhan AN, Penland L, Sinha R, Gillich A, Sit RV, Chang S, Conley SD, Mori Y, Seita J, Berry GJ, Shrager JB, Metzger RJ, Kuo CS, Neff N, Weissman IL, Quake SR, Krasnow MA. A molecular cell atlas of the human lung from single-cell RNA sequencing. Nature 2020;587:619–625.

35. O’Dwyer DN, Zhou X, Wilke CA, Xia M, Falkowski NR, Norman KC, Arnold KB, Huffnagle GB, Murray S, Erb-Downward JR, Yanik GA, Moore BB, Dickson RP. Lung Dysbiosis, Inflammation, and Injury in Hematopoietic Cell Transplantation. AmJRespirCritCare Med 2018;198:1312–1321.

36. O’Dwyer DN, Ashley SL, Gurczynski SJ, Xia M, Wilke C, Falkowski NR, Norman KC, Arnold KB, Huffnagle GB, Salisbury ML, Han MK, Flaherty KR, White ES, Martinez FJ, Erb-Downward JR, Murray S, Moore BB, Dickson RP. Lung Microbiota Contribute to Pulmonary Inflammation and Disease Progression in Pulmonary Fibrosis. Am J Respir Crit Care Med 2019;199:1127–1138.

37. Peled JU, Gomes ALC, Devlin SM, Littmann ER, Taur Y, Sung AD, Weber D, Hashimoto D, Slingerland AE, Slingerland JB, Maloy M, Clurman AG, Stein-Thoeringer CK, Markey KA, Docampo MD, Burgos da Silva M, Khan N, Gessner A, Messina JA, Romero K, Lew MV, Bush A, Bohannon L, Brereton DG, Fontana E, Amoretti LA, Wright RJ, Armijo GK, Shono Y, et al. Microbiota as Predictor of Mortality in Allogeneic Hematopoietic-Cell Transplantation. NEnglJMed 2020;382:822–834.

38. Abreu NA, Nagalingam NA, Song Y, Roediger FC, Pletcher SD, Goldberg AN, Lynch SV. Sinus microbiome diversity depletion and Corynebacterium tuberculostearicum enrichment mediates rhinosinusitis. SciTranslMed 2012;4:151ra124.

39. Kim H-S, Arellano K, Park H, Todorov SD, Kim B, Kang H, Park YJ, Suh DH, Jung ES, Ji Y, Holzapfel WH. Assessment of the safety and anti-inflammatory effects of three Bacillus strains in the respiratory tract. Environ Microbiol 2021;doi:10.1111/1462-2920.15530.

40. Nieto A, Mazón A, Nieto M, Calderón R, Calaforra S, Selva B, Uixera S, Palao MJ, Brandi P, Conejero L, Saz-Leal P, Fernández-Pérez C, Sancho D, Subiza JL, Casanovas M. Bacterial Mucosal Immunotherapy with MV130 Prevents Recurrent Wheezing in Children: A Randomized, Double-Blind, Placebo-controlled Clinical Trial. Am J Respir Crit Care Med 2021;204:462–472.

41. Garcia O, Hiatt MJ, Lundin A, Lee J, Reddy R, Navarro S, Kikuchi A, Driscoll B. Targeted Type 2 Alveolar Cell Depletion. A Dynamic Functional Model for Lung Injury Repair. Am J Respir Cell Mol Biol 2016;54:319–330.

42. Dahmer MK, Flori H, Sapru A, Kohne J, Weeks HM, Curley MAQ, Matthay MA, Quasney MW, BALI and RESTORE Study Investigators and Pediatric Acute Lung Injury and Sepsis Investigators (PALISI) Network. Surfactant Protein D Is Associated With Severe Pediatric ARDS, Prolonged Ventilation, and Death in Children With Acute Respiratory Failure. Chest 2020;158:1027–1035.

43. Distler JHW, Györfi A-H, Ramanujam M, Whitfield ML, Königshoff M, Lafyatis R. Shared and distinct mechanisms of fibrosis. Nat Rev Rheumatol 2019;15:705–730.

44. Crosby LM, Waters CM. Epithelial repair mechanisms in the lung. Am J Physiol Lung Cell Mol Physiol 2010;298:L715–731.

45. Hu B, Liu J, Wu Z, Liu T, Ullenbruch MR, Ding L, Henke CA, Bitterman PB, Phan SH. Reemergence of hedgehog mediates epithelial-mesenchymal crosstalk in pulmonary fibrosis. Am J Respir Cell Mol Biol 2015;52:418–428.

46. Didiasova M, Singh R, Wilhelm J, Kwapiszewska G, Wujak L, Zakrzewicz D, Schaefer L, Markart P, Seeger W, Lauth M, Wygrecka M. Pirfenidone exerts antifibrotic effects through inhibition of GLI transcription factors. FASEB J 2017;31:1916–1928.

47. Malaviya R, Kipen HM, Businaro R, Laskin JD, Laskin DL. Pulmonary toxicants and fibrosis: innate and adaptive immune mechanisms. Toxicol Appl Pharmacol 2020;409:115272.

48. Haupt S, Caramia F, Klein SL, Rubin JB, Haupt Y. Sex disparities matter in cancer development and therapy. Nat Rev Cancer 2021;doi:10.1038/s41568-021-00348-y.

49. Wagner AD, Grothey A, Andre T, Dixon JG, Wolmark N, Haller DG, Allegra CJ, de Gramont A, VanCutsem E, Alberts SR, George TJ, O’Connell MJ, Twelves C, Taieb J, Saltz LB, Blanke CD, Francini E, Kerr R, Yothers G, Seitz JF, Marsoni S, Goldberg RM, Shi Q. Sex and Adverse Events of Adjuvant Chemotherapy in Colon Cancer: An Analysis of 34□640 Patients in the ACCENT Database. J Natl Cancer Inst 2021;113:400–407.

50. Diefenhardt M, Ludmir EB, Hofheinz R-D, Ghadimi M, Minsky BD, Rödel C, Fokas E. Association of Sex With Toxic Effects, Treatment Adherence, and Oncologic Outcomes in the CAO/ARO/AIO-94 and CAO/ARO/AIO-04 Phase 3 Randomized Clinical Trials of Rectal Cancer. JAMA Oncol 2020;6:294–296.

