## Supplemental Data for "Metatranscriptomic Signatures of Lung Function in Pediatric Hematopoietic Cell Transplant Candidates"

**Online Data Supplement**

**Contents:**

1. Supplemental Methods 1) Clinical Protocols
2. Supplemental Methods 2) Pulmonary Function Testing
3. Supplemental Methods 3) Laboratory Protocols
4. Supplemental Methods 4) Metatranscriptome Protocols
5. Supplemental Methods 5) Data Analysis
6. Table E1) Classification of Abnormal Pulmonary Function Tests
7. Figure E1) Spearman Correlation Matrix for PFT Measurements
8. Figure E2) Cumulative Incidence of Fatal Post-HCT Lung Injury
9. Figure E3) Spearman Correlation Matrix for Pulmonary Microbes
10. Figure E4) BAL Microbiome Principal Component Analysis
11. Figure E5) Association between BAL Microbiome and DLCO/Va
12. Supplemental References

**Supplemental Methods 1) Clinical Protocols**

***Patients:*** A universal screening protocol to evaluate pulmonary health prior to pediatric HCT was instituted at the University Medical Center in Utrecht, the Netherlands, starting in 2007. BAL was performed in all pediatric HCT candidates as standard-of-care routine clinical practice in order to identify potential pulmonary pathogens and reduce the risk of post-HCT lung injury. BAL was only performed in children undergoing general anesthesia for other indications; therefore, there was a small subset of patients who did not require general anesthesia and therefore could not contribute clinical samples (1). Patients were enrolled with informed consent under the University Medical Center, Utrecht, Institutional Ethics Committee approval numbers 05/143 and 11/063.

***BAL Specimen Collection:*** Standard clinical practice for pediatric bronchoscopy involved endotracheal intubation followed by instillation of 10 mL of normal saline aliquots through an endotracheal catheter wedged in the distal bronchi. After aliquoting for clinical testing, remaining BAL was stored at -70°C for future research. Most samples were thawed zero or one times prior to processing for this study.

***Clinical Protocols:*** The following clinical protocols are discussed to contextualize care received by study patients and the approach utilized to reduce post-HCT mortality. First, details of clinical microbiologic testing strategy have been previously described (1–4). Approach to modifying clinical care based on pre-HCT BAL clinical microbiology test results is described in the same references and entails addition or modification of immunosuppression and/or antimicrobials in high-risk patients and/or those with new pulmonary symptoms. HCT conditioning regimens have been described in detail previously and broadly entail use of busulfan and fludarabine for patients with non-malignant disease and TBI with etoposide or busulfine and fludarabine or clofarabine with fludarabine for patients with malignant diseases (1). Serotherapy (typically ATG) was used for unrelated donor transplants; this was excluded for high-risk malignancies receiving UCB starting in December 2012. GVHD prophylaxis incorporated cyclosporine with prednisolone in UCB recipients and methotrexate in unrelated donor transplants (all study years) and HLA-matched siblings (starting in 2013). Antimicrobial prophylaxis included ciprofloxacin and fluconazole through neutropenia with the addition of cefazolin during mucositis; co-trimoxazole three times weekly starting on day +28 for *Pneumocystis* prophylaxis; acyclovir if pre-HCT HSV seropositive or if VZV seropositive and receiving umbilical cord blood (UCB); and voriconazole (2005-2015) or amphotericin B (2015-present) if at high-risk for IFI. Febrile neutropenia was treated empirically with ceftazidime and vancomycin. Treatment for alloreactive lung injury involved intravenous methylprednisolone (10 mg/kg/day) for 3 days followed by 2 mg/kg/day with weekly 25% tapers if stabilization/improvement or up to 6 monthly methylprednisolone pulses for refractory disease; azithromycin was also used for immunomodulatory effect.

***Outcomes:*** All-cause mortality was documented in all patients through 2 years post-HCT with no loss to follow-up. Post-HCT lung injury was defined as lower respiratory tract infection requiring supplemental oxygen or alloreactive lung syndromes as further defined by the American Thoracic Society and National Institutes of Health chronic GVHD criteria for Idiopathic Pneumonia Syndrome and Bronchiolitis Obliterans respectively (5, 6).

**Supplemental Methods 2) Pulmonary Function Testing**

**Acquisition of Pulmonary Function Testing:** PFTs included spirometry, whole body plethysmography, and measurement of carbon monoxide diffusion capacity and were performed in developmentally capable children ages 4 and older according to American Thoracic Society / European Respiratory Society criteria used a calibrated pneumotachometer system (Jaeger, Hochberg, Germany) (7, 8). Patients who could not perform spirometry adequately either due to developmental age or pulmonary function were not included in this study. Some patients who performed spirometry could not perform plethysmography or diffusion testing. Pre-bronchodilator PFT values were normalized for age, height, sex, and race and expressed as (1) a percentage deviation from the mean (% predicted) according to healthy Dutch children and (2) a standardized deviation from the mean (Z-score) according to the Global Lung Index (GLI) (9, 10). Z-score was not evaluable in 2 patients due to medical chart documentation of only the % predicted values and not the absolute values.

**Interpretation of PFTs:** Discrimination of normal vs abnormal values necessarily requires dichotomization of the cohort around a threshold. Therefore, we tested two approaches to patient classification. ***Standard classification:*** we defined obstruction (FEV_1_/FVC absolute ratio < 0.80), restriction (TLC_%pred_ <80% or if plethysmography not available, both FVC_%pred_ and FEV1_%pred_ <80%), mixed spirometry (meets criteria for both obstruction and restriction), diffusion impairment (DLCO/Va_%pred_ <80%), and air trapping (RV/TLC absolute ratio > 0.25). ***GLI-recommended classification:*** we defined obstruction (FEV_1_/FVC_z-score_ <-1.64), restriction (TLC_z-score_ <-1.64 or if plethysmography not available, both FVC_z-score_ and FEV1_z-score_ <-1.64), mixed (meets criteria for both obstruction and restriction), diffusion impairment (DLCO/Va_z-score_ <-1.64), and air trapping (RV/TLC_z-score_ >1.64). Z-score cutoffs of ±1.64 were chosen since they reflect the upper/lower 5% of the population (upper limit of normal, ULN; lower limit of normal, LLN) (7, 10). Comparison of two classification approaches is presented in **Supplemental Table 1**.

**Supplemental Methods 3) Laboratory Protocols**

***BAL RNA Extraction:*** Samples underwent a previously described RNA extraction protocol optimized for BAL fluid (11). 200uL of BAL was combined with 200uL DNA/RNA Shield (Zymo) and 0.5mm glass bashing beads (Omni) for 5 cycles of 25 seconds bashing at 30Hz with 60 seconds of rest on ice between each cycle (TissueLyser II, Qiagen). Subsequently, samples were centrifuged for 10 minutes at 4°C and the supernatant was used for column-based RNA extraction with DNase treatment according to the manufacturer’s recommendations (Zymo ZR-Duet DNA/RNA MiniPrep Kit). Resultant RNA was eluted in 5uL sterile water and stored at -70°C until sequencing library preparation.

***Sequencing Library Preparation:*** Samples underwent a previously described sequencing library preparation protocol optimized for BAL fluid (12). First, BAL RNA was dehydrated at 40°C for 25 minutes in a 384 well plate (GeneVac E-Z2). Second, sequencing libraries were prepared using miniaturized protocols adapted from the NEB Ultra II RNA Library Prep Kit (Step-by-step protocol is published here <https://www.protocols.io/private/07DC2C6765A0D9665C227879D935EED3>). Reagents were dispensed using the Echo 525 (Labcyte) and underwent Ampure-XP bead cleaning on a Beckman Coulter Biomek NX^P^ instrument. Libraries underwent 19 cycles of PCR amplification, size selection to a target 300-700 nt, and were pooled to facilitate approximately even depth of sequencing.

***Positive and Negative Controls:*** 25 picograms (pg) of External RNA Controls Consortium (ERCC) pooled standards were spiked-in to each sample after RNA extraction and before library preparation to serve as internal positive controls (Thermo Fisher Scientific Cat. No 4456740). In addition, to identify contamination in laboratory reagents and the laboratory environment, we performed RNA extraction and sequencing on 2 samples of 200uL sterile water and 2 samples of 200uL HeLa cells taken from a laboratory stock. These samples were processed at the same time as the patient BALs in order to use the same lot of reagents and minimize batch effect on control samples.

***Sequencing Protocol:*** Samples were pooled across 4 lanes of an Illumina NovaSeq 6000 instrument and sequenced to a target depth of 40 million read-pairs with sequencing read length of 125 nt.

**Supplemental Methods 4) Metatranscriptome Protocols**

***Processing of .fastq Sequencing Files:*** All sequencing files were processed using the IDseq pipeline v3.13 (<https://github.com/chanzuckerberg/idseq-web>) (13). Briefly, .fastq files underwent a first round of human read subtraction (STAR) followed by Illumina adaptor removal (Trimmomatic), quality filtering (PriceSeq), duplicate read removal (CD-HIT-DUP), and LZW complexity filtering. Next, sequencing files underwent a second more stringent round of human read subtraction (Bowtie2) followed by a third round of human read subtraction (STAR), subsampling to 1 million fragments, and a fourth and final round of human read subtraction (GSNAP). Resultant sequencing files underwent alignment to the NCBI nt/nr database using GSNAP with minimum alignment length >36. Taxa counts were generated with associated metrics of percent identity, contig length, and e-value. Quality metrics for the sequencing run including percent of reads that passed the PriceSeq filter step and percent of reads that passed all steps were examined and samples with poor sequencing quality were re-sequenced.

***Quality Control:*** Taxa count files were generated for each sample; taxa not detected were determined to have a count of zero. To reduce spurious associations due to ambiguous alignments, counts with taxa alignment identities <85% were replaced with zero. ERCC scatterplots of pg input vs. sequencing reads output were generated and analyzed for each sample to ensure the positive control (ERCC spike-in) was adequately detected and that each of the 92 transcripts within the ERCC spike-in were proportional to their original input concentrations (suggesting linear sequencing). Subsequently, original RNA mass of the sample was back-calculated by solving the linear proportionality equation (total sample reads / total sample mass) = (ERCC reads / ERCC mass), where sample reads and ERCC reads were detected by the above protocol and ERCC input was standardized as 25 picograms. The calculated sample mass was then reduced by 25pg (the ERCC input) to equal the original sample mass before ERCC addition. Since the input RNA mass of the water controls was determined to be ~5 pg (presumably reflecting 5 pg of sequenceable contamination), we discarded samples whose total input mass was <10 pg as we were unable to reliably differentiate between contamination and true constituents.

***Calculation of Sample x Taxa Mass Matrices:*** Matrices of taxa mass as calculated above were generated for every taxa detected in every sample. Taxa not detected in any sample were determined to be below the lower limit of detection (1 unique read passing all quality control filters designated above) and were set as 0 pg in value. Next, Sample x Taxa Mass matrices were tested for contamination as follows. ***Correction for Contamination:*** Here we relied on the observation that contaminant taxa are PCR amplified to a greater extent in low biomass samples undergoing library preparation, and hence are overrepresented in sequencing data of low biomass samples (14, 15). Therefore, for every taxa detected in the cohort, we tested for an inverse relationship between log_10_-transformed sequencing reads and original pre-PCR log_10_-transformed sample mass by fitting linear regression models. Taxa that could be modeled with good fit (adjusted R^2^≥0.6) were determined to be contaminants. We then calculated contamination-adjusted taxa counts by adjusting for an inflation factor equal to the linear regression coefficient (slope) as follows: *Log_10_(adjusted taxa reads) = Log_10_(original taxa reads) + (regression slope)* Log_10_(sample RNA mass).* Finally, contaminant-adjusted reads were used to calculate the original mass of each taxa in each sample by solving the ERCC linear proportionality equation as listed above. ***Correction for Batch Effect:*** Samples were extracted in 27 batches and underwent library preparation in 2 batches. For each contaminating taxa listed in the table above, we tested for batch effect by plotting the taxa mass relative to the sample processing order. We identified a natural breakpoint after batch 9 where for batches 10-27, the mass of *Acinetobacter*, *Aureobasidium*, *Rhodotorula*, and *Scedosporium* increased by 2 orders of magnitude with the introduction of new reagents. To correct for this, for all samples in batches 10-27, the mass of these 4 taxa was reduced by the difference in means between the batches 1-9 vs. 10-27. There were no batch effects related to taxa mass detected across the 2 library preparation batches.

***Calculation of Sample Microbiome Traits:*** Total bacterial, viral, and fungal mass were calculated using the sum of all taxa in those kingdoms that could be identified at the genus level. Reads aligning to bacteria at the taxonomic level of family that could not be assigned to the taxonomic level of genus were not included in this calculation. ***Alpha Diversity*** was calculated as the *Simpson’s Alpha Diversity Index = 1 – (∑ [microbe mass]*[microbe mass – 1] / ([total microbial mass] * [total microbial mass – 1])* where microbe mass was calculated for each species present in each sample. Alpha diversity was calculated for all species (total microbial diversity) and again for bacterial total alone. ***Richness*** was calculated as the number of species occupying ≥1% of the total microbial mass. ***Microbial Dominance*** was calculated as the percent of the total microbial mass occupied by the most abundant taxa in each sample.

***Calculation of Sample x Gene Expression Matrices:*** Sequencing files were aligned to human genome v38 using STAR as described above in the IDSeq pipeline (<https://github.com/chanzuckerberg/idseq-web>). HFNC Symbols were matched to ENSEMBL IDs using *biomart*. Non-protein coding genes were removed. Protein coding genes that were not identified in at least 25% of samples were removed. Samples that did not have at least 50,000 total counts to protein-coding genes were removed.

**Supplemental Methods 5) Data Analysis**

***Correlation between Individual PFTs:*** Pair-wise Spearman rank-based correlation was calculated for each PFT pair using complete observations (e.g.: FVC_%pred_ and FEV1_%pred_, FVC_%pred_ and TLC_%pred_, etc) with FDR correction for multiple comparisons. Spearman correlation values were plotted in a heatmap with correlation-based clustering of rows and columns to identify clusters of highly-correlated PFTs (**Supplemental Figure 1**).

***Correlation between PFTs and Clinical Characteristics:*** Correlation between each PFT and patient age was tested using Spearman rank-based correlation. Correlation between each PFT and patient sex was tested using Wilcoxon rank-sum testing. Correlation between each PFT and HCT indication (categorized into 5 groups) was tested using the Kruskal-Wallis test. All analyses underwent false discovery rate (FDR) adjustment for multiple comparisons.

***Association between PFTs and Post-HCT Outcomes:*** Time-dependent survival estimates for all-cause mortality were calculated using Kaplan-Meier function (*survival* package). Differences in overall survival estimates were tested using Cox proportional hazards models for continuous variables (e.g. PFT measurements). The time-dependent cumulative incidence function for fatal lung injury were calculated using relapse and non-lung injury related mortality as competing risks (*cmprsk* package). Differences in cumulative incidence of these outcomes were assessed across continuous variables (e.g. PFT values) using Fine & Gray’s test and the subdistribution hazard ratio (SHR).

***Association between Individual BAL Microbes:*** First, to reduce spurious low-abundance taxa, genus-level sample x taxa mass matrices were subset to include only genera present at ≥ 0.25 fg in ≥ 50% of the cohort. Next, the mass for microbes that were undetectable in any given sample was imputed using Bayesian posterior estimates to approximate the lower limit of detection in a given sample (*cmultRepl).* The sample x taxa mass matrix then underwent variance stabilizing transformation (*vst*, *DESeq2*) and the taxa-taxa Spearman correlation matrix was constructed using Euclidean-distance clustering of rows and columns to identify highly correlated sets of genera.

***BAL Microbe Principal Component Analysis:*** Genus-level sample x taxa mass matrices were subset to include only genera present at ≥ 0.25 fg in ≥ 50% of the cohort. The centered and scaled matrix was using in PCA (*prcomp*) and the relationship between microbes and principal components 1 and 2 was plotted. The values of each patient BAL along PC1 and 2 were then extracted and tested for association with each PFT using Spearman rank-based correlation with FDR adjustment for multiple comparison.

***Association between PFTs and BAL Microbiome Traits:*** We tested for association between each PFT and total bacterial, viral, and fungal mass, total mass of major bacterial phyla, alpha diversity, richness, and dominance using Spearman rank correlations with FDR adjustment for multiple comparisons.

***Association between PFTs and BAL Microbes (Differential Abundance):*** To allow analysis of rarer genera, we subset the taxa matrix to include genera present at ≥ 0.2 fg in ≥ 25% of the cohort. We then estimated the negative binomial dispersion of each genus in the population and associated each microbial genus with each PFT using generalized linear models with likelihood ratio tests again applying FDR adjustment for multiple comparison (*edgeR*). Taxa showing positive or negative association with selected PFTs were graphically depicted using volcano plots. Since we earlier identified that patient age and sex were also associated with PFTs, we repeated the analysis adjusting for these covariates in multivariable models to test if microbe-PFT associations could be demonstrated independent of age and sex.

***Association between PFTs and MSigDB Hallmark Gene Expression:*** The Molecular Signatures Database Hallmark Gene set consists of 50 curated gene sets representing pathways fundamental to human biology. For each BAL, we calculated gene set enrichment scores for each of the n=50 gene sets using Poisson distributions (*gsva,* ***figure below***). We then associated gene set enrichment scores with PFT results using Spearman rank based correlation and plotted the results in a heatmap with clustering of rows and columns based on Euclidean distance in order to identify correlations between PFTs and hallmark gene sets.

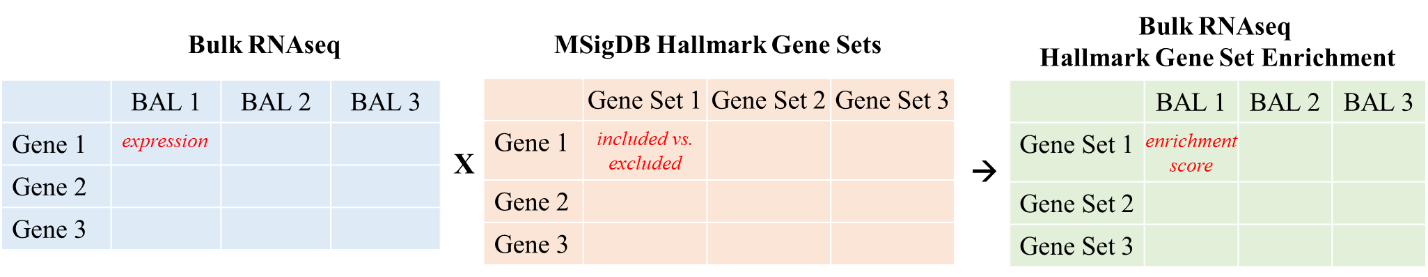

***Association between PFTs and BAL Gene Expression (Differential Expression):*** To test for individual genes whose expression was associated with PFT measurements, we estimated the negative binomial dispersion of each gene in the population and associated each gene distribution with each PFT using generalized linear models with likelihood ratio tests again applying FDR adjustment for multiple comparison (*edgeR*). Since we earlier identified that patient age and sex were also associated with PFTs, we repeated the analysis adjusting for these covariates in multivariable models to test if transcript-PFT associations could be demonstrated independent of age and sex. Shared ontology was tested using pathway analysis (*enrichR*).

***Imputation of BAL Cell Fractions:*** Recent advances in single cell sequencing techniques have facilitated development of robust organ-specific cellular atlases enumerating greater cell types with greater specificity than previously possible. To impute BAL cell type fractions, we applied a lung single cell reference atlas produced by *Travaglini et al, Nature 2020* to our bulk sequencing data. A matrix of gene counts per single cell (n=9409 cells) was subset to include protein coding genes and used to generate a reference of cell-type specific marker genes using CiberSortX (signature matrix, run on Linux server through Docker) (16). For this analysis, 58 provided cell types were collapsed into 15 cell types based on shared ontology and after removing stromal and sub-epithelial cells not likely to be captured in lavage fluid. Next, the signature matrix of cell type-specific marker genes was applied to the matrix of gene counts per BAL sample to estimate cell type fractions within each sample (***figure*** ***below***). Highly correlated cell types were identified using Spearman rank based correlation with hierarchical clustering.

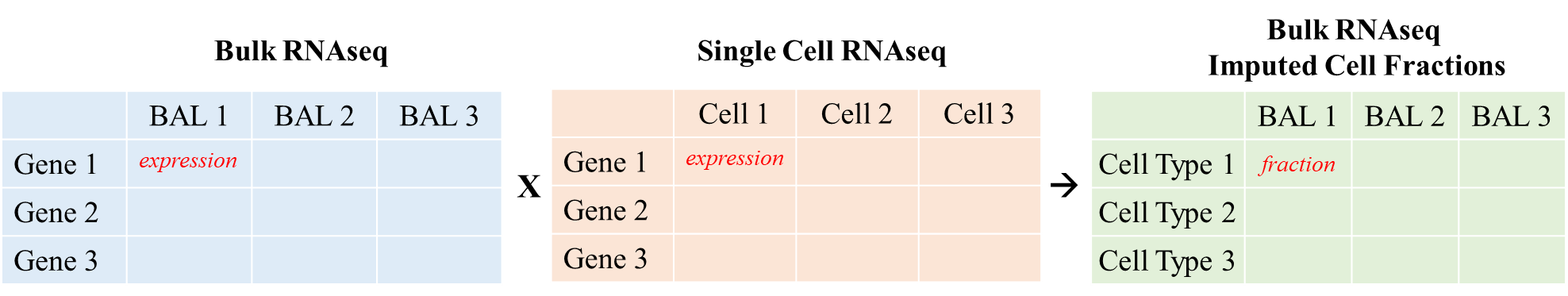

***Imputation of BAL Cell Type Specific Gene Expression:*** Using the signature matrix of cell type-specific markers described above, 15 cell types were further collapsed into 7 overarching cell types based on shared ontology and then average gene expression matrices for each BAL cell type were imputed first for patients with pulmonary restriction and then for patients without pulmonary restriction (CiberSortX group mode) (16). The result of this step is two matrices (one for patients with restriction and one for patients without restriction) composed of genes (rows) x cell types (columns) where cell value is mean ± standard error expression (***figure*** ***below***). To test for differences in average cell type-specific gene expression across these two groups (restriction vs no restriction), Welch's T-test of gene expression means with unequal variances was applied to each gene for each cell type across the 2 groups, with FDR adjustment for multiple comparisons.

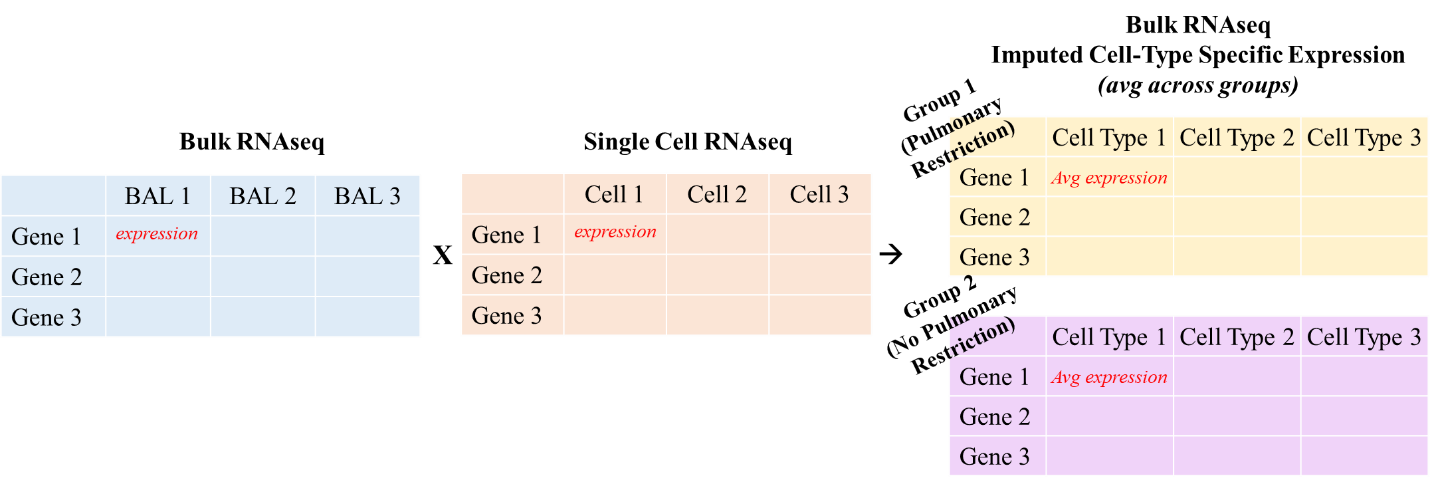
Next, again using the signature matrix of cell type-specific markers described above for 7 overarching cell types, cell-type specific gene expression was imputed for each patient. The result of this step is 7 matrices (one for each cell type) composed of genes (rows) x patients (columns) where cell value is mean expression (***figure*** ***below***). To test for associations between cell type-specific gene expression and pulmonary function tests (e.g.: FVC_% predicted_), we fit negative binomial generalized linear models (NB-GLM, *edgeR*) with adjustment for false discovery.

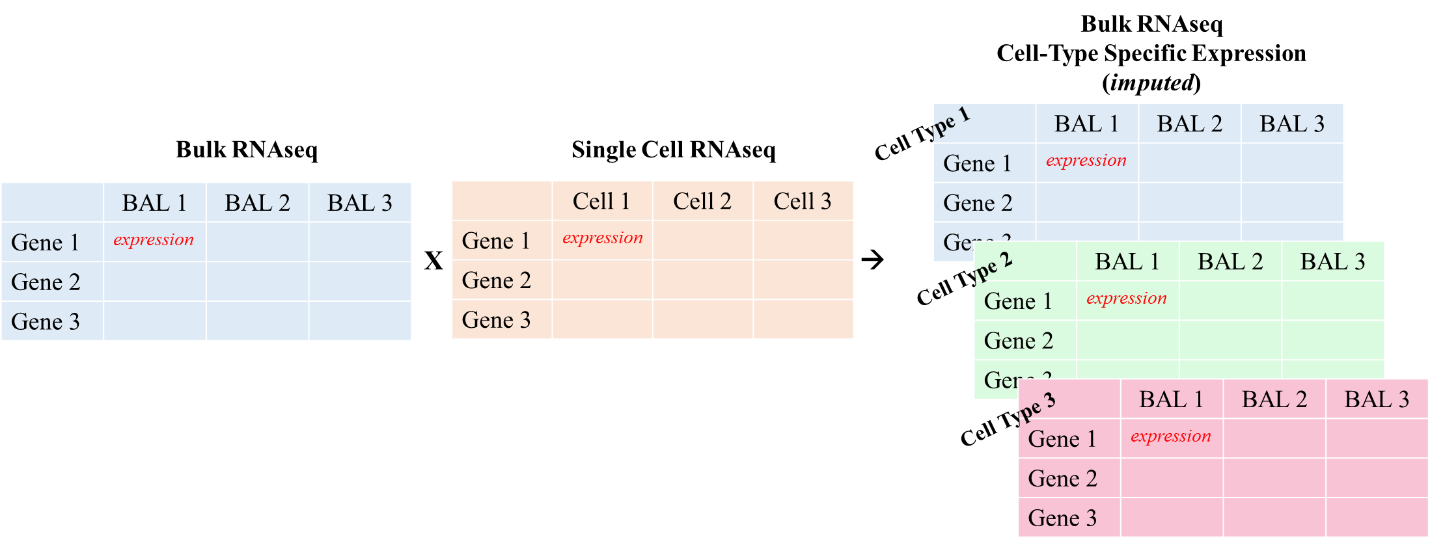

***Association Between BAL Metatranscriptome, PFTs, and Post-HCT Outcomes:*** We tested for associations between taxa mass and post-HCT mortality using NB-GLM (*edgeR*) with adjustment for FVC_%pred_, age, and sex. Interaction terms were added to test for taxa*PFT interactions with respect to outcome. We tested for associations between Hallmark Gene Set enrichment scores and post-HCT mortality using Cox regression models with adjustment for FVC_%pred_, age, and sex. We then tested for improvement of a Cox regression model containing FVC_%pred_, age, and sex after adding Hallmark Apical Surface gene set enrichment score and BAL *Haemophilus* mass (vst-transformed). Comparison of the two models was assessed using the likelihood ratio test. We then plotted estimated survival curves according to the Cox regression model at specified values of FVC_%pred_, Haemophilus mass, and Hallmark Apical Surface gene set enrichment.

**Table E1: Classification of Abnormal Pulmonary Function Tests**

|  | **Classification 1 - Standard** | **Classification 2 – GLI Recommended** |
| --- | --- | --- |
| **Obstruction** | FEV_1_/FVC < 0.80  n=12/104 | FEV_1_/FVC_z-score_ <-1.64  n=7/104 |
| **Restriction** | TLC_%pred_ <80%  *or if plethysmography not available, both FVC_%pred_ and FEV1_%pred_ <80%*  n=35/104 | TLC_z-score_ <-1.64  *or if plethysmography not available, both FVC_z-score_ and FEV1_z-score_ <-1.64*  n=29/104 |
| **Mixed** | meets criteria for both obstruction and restriction  n=1/104 | meets criteria for both obstruction and restriction  n=0/104 |
| **Diffusion Impairment** | DLCO/Va_%pred_ <80%  n=21/63 | DLCO/Va_z-score_ <-1.64  n=14/63 |
| **Air Trapping** | RV/TLC > 0.25  n=44/78 | RV/TLC_z-score_ >1.64  n=16/78 |
| **Any Abnormality** | ≥1 of the above 5  n=80/104 | ≥1 of the above 5  n=54/104 |

**Legend:** PFTs were classified into obstruction, restriction, or mixed, with/without diffusion impairment, with/without air trapping, per two classification schema listed above.

**Figure E1) Spearman Correlation Matrix for PFT Measurements**

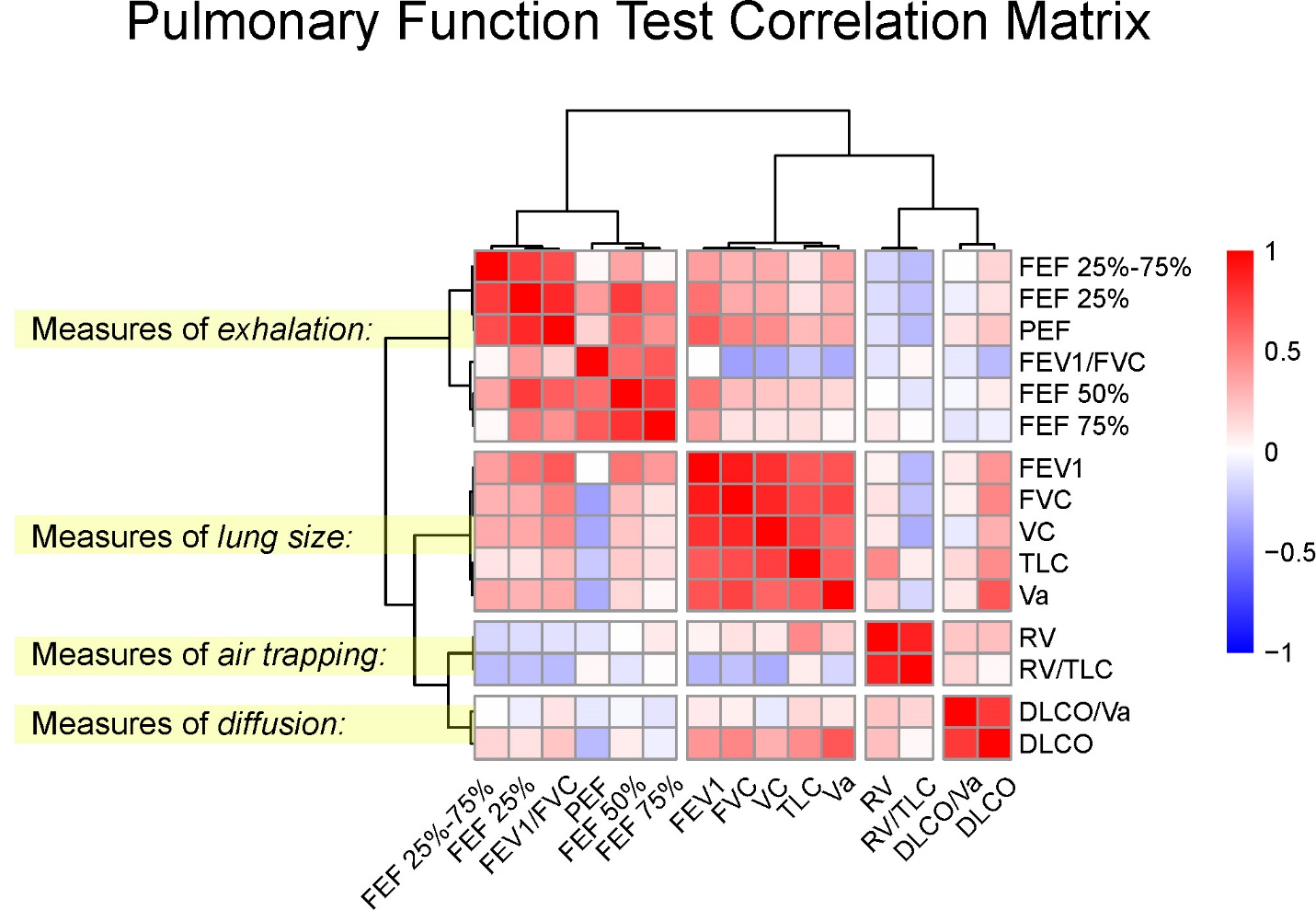

**Legend:** Spearman correlation using pairwise complete observations of PFTs expressed as percent of predicted with columns and rows clustered using correlation distances. Four clusters are identified corresponding to measures of exhalation, lung size, air trapping, and diffusion.

**Figure E2) Cumulative Incidence of Early Fatal Post-HCT Lung Injury**

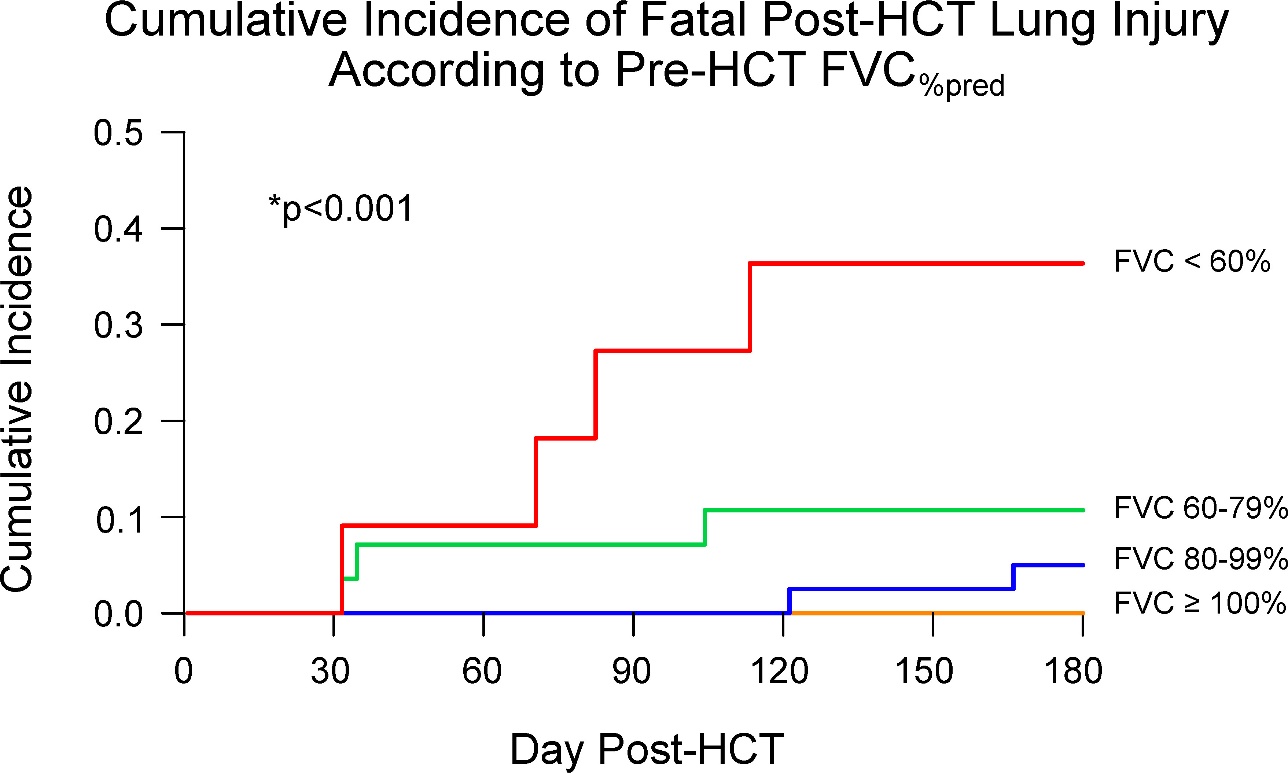

**Legend:** Cumulative incidence of post-HCT fatal lung injury in the first 6 months according to pre-HCT FVC_%pred_. Worse FVC_%pred_ was associated with greater cumulative incidence of fatal post-HCT lung injury in the first 6 months post-HCT after accounting for relapse and death without lung injury as a competing risk (competing risk regression p<0.001).

**Figure E3) Spearman Correlation Matrix for Pulmonary Microbes**

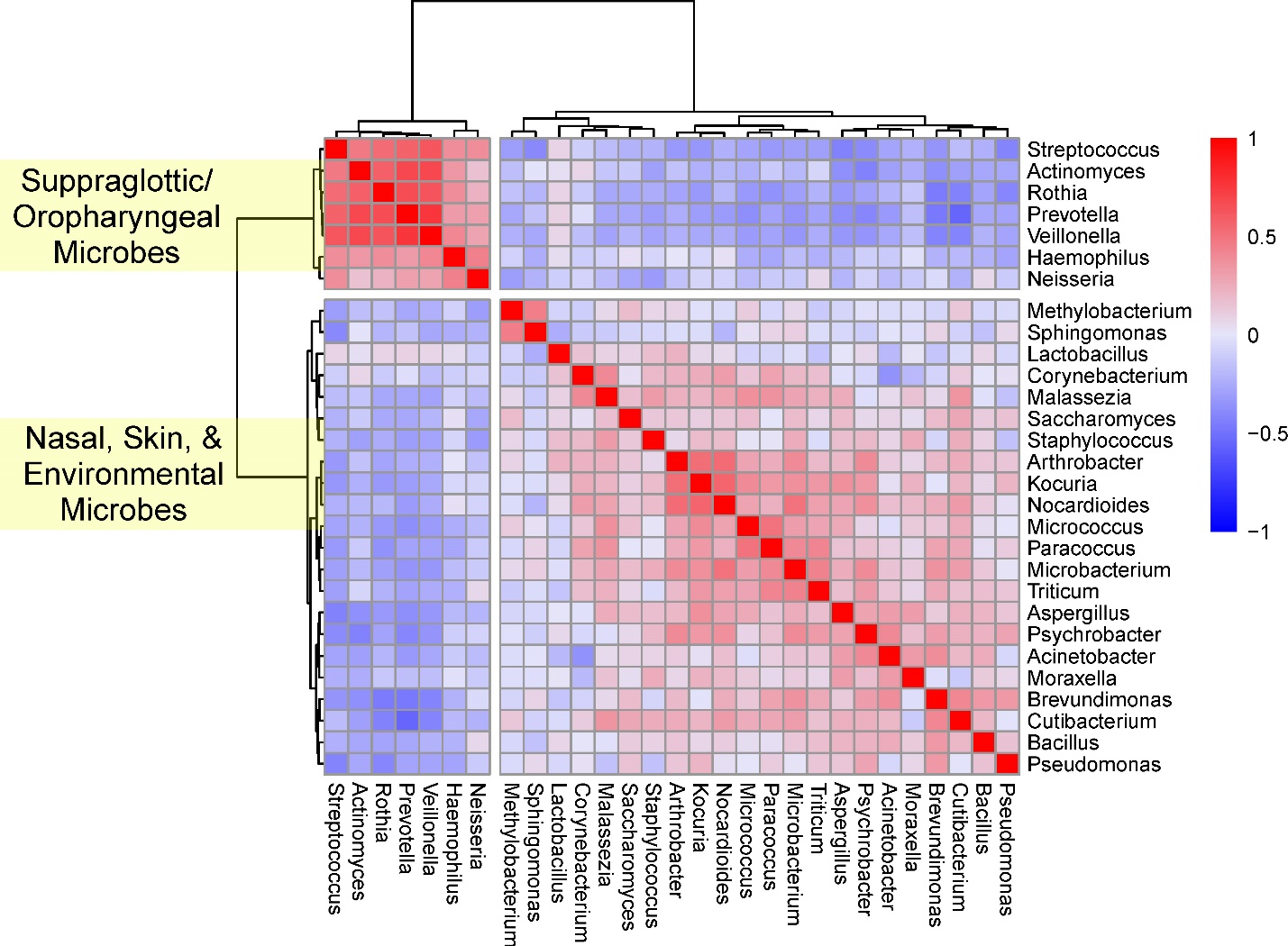

**Legend:** Microbial genera were first subset to include only taxa present in in ≥50% of samples at a minimum mass of 0.25pg. Taxa then underwent variance-stabilizing transformation (*vst*). Spearman correlation using pairwise complete observations with columns and rows clustered using Euclidean distances.

**Figure E4) BAL Microbiome Principal Component Analysis**

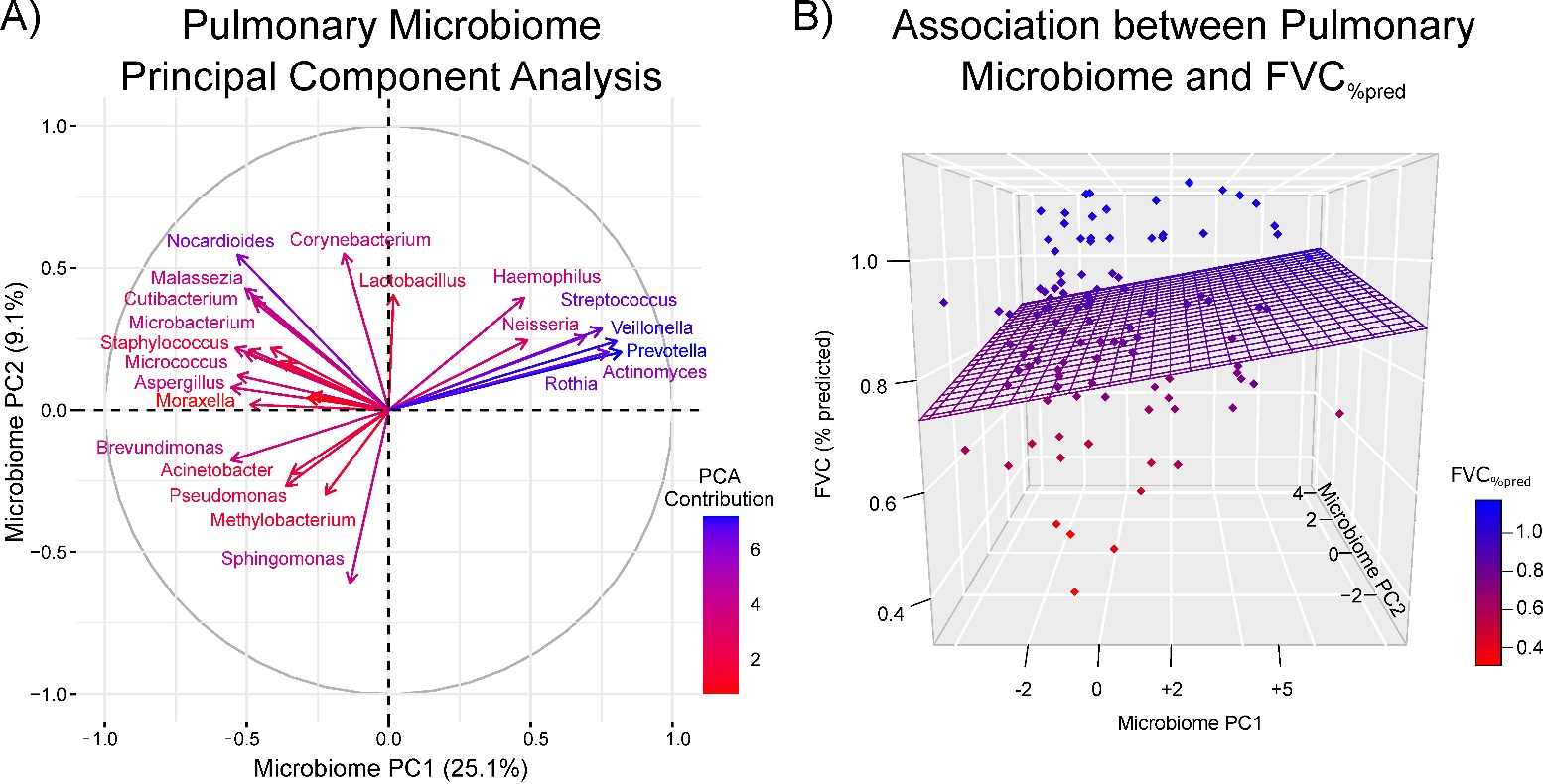

**Legend: (A)** Principal component analysis of BAL microbes present in ≥50% of samples at a minimum mass of 0.25pg. Microbial masses underwent variance stabilization transformation (*vst*), centering, and scaling prior to PCA. **(B)** 3-dimensional scatter plot relating FVC and BAL microbiome. Each patient is represented as a single dot plotted in 3 dimensions according to the patient’s measured FVC_%pred_ (y-axis), the patient’s BAL coordinates along Principal Component 1 of the PCA generated in Figure 2a (x-axis), and the patient’s BAL coordinates along Principal Component 2 of the PCA generated in Figure 2a (z-axis). Dot shading correlates with FVC_%pred_. The grid consists of estimated values of FVC_%pred_ (z-axis) according to a linear Gaussian model using BAL principal components 1 (x-axis) and 2 (z-axis). The tilting of the grid towards the front-left of the 3D box shows that lower values along PC1 and 2 are associated with lower FVC_%pred_ whereas the tilting of the grid towards the back-right of the 3D box higher values of PC1 and 2 are associated with higher FVC_%pred_.

**Figure E5) Association between BAL Microbiome and DLCO/Va**

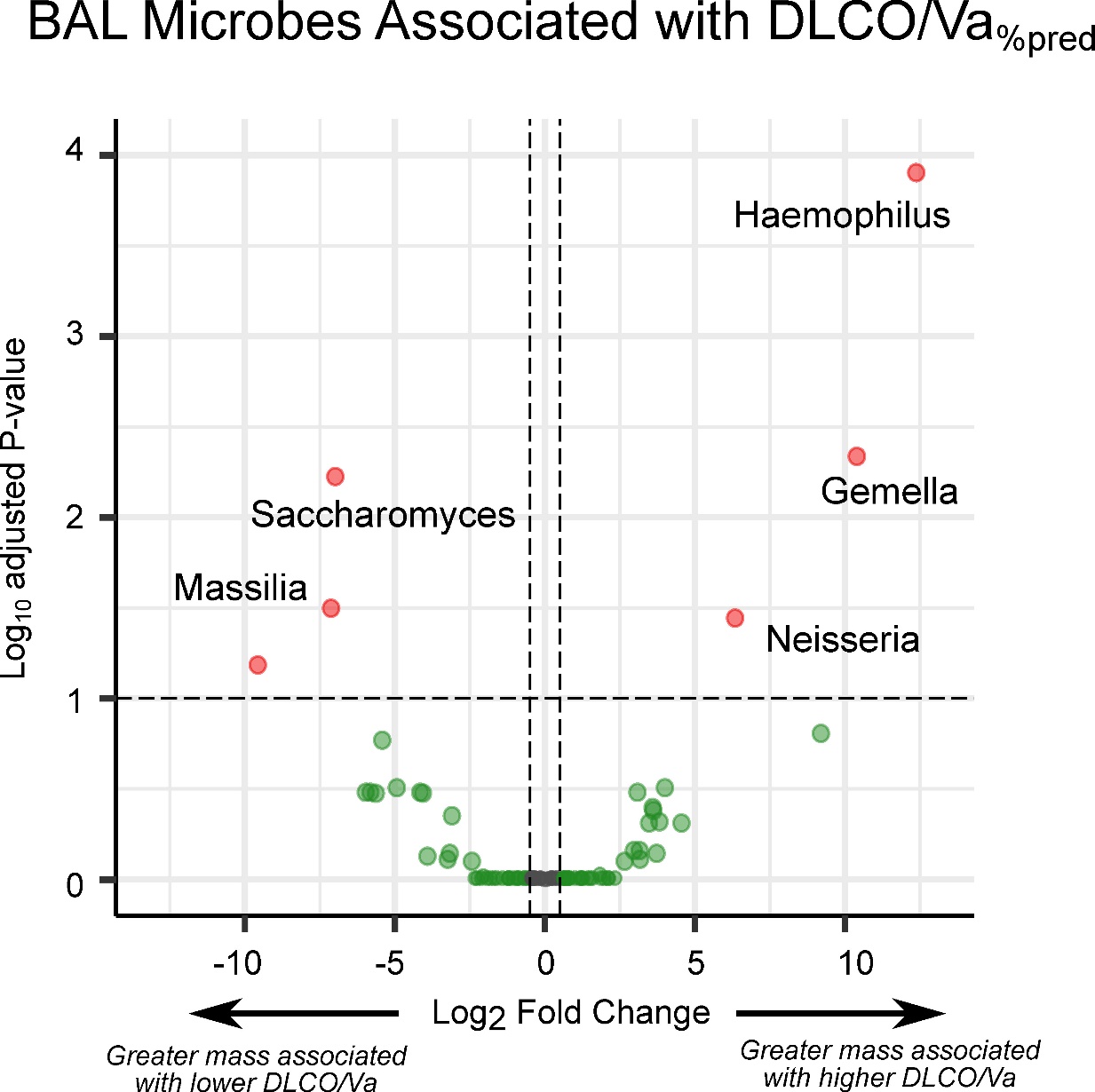

**Legend:** Volcano plot showing the association between BAL microbe mass and DLCO/Va_%pred_. Associations tested using negative binomial generalized linear models accounting for patient age and sex covariates. Positive log_2_ fold change (x-axis) indicates that greater microbial mass is associated with greater DLCO/Va _%pred_; negative log_2_ fold change indicates that greater microbial mass is associated with lower DLCO/Va _%pred_.

**References**

E1.	Versluys B, Bierings M, Murk JL, Wolfs T, Lindemans C, Vd Ent K, Boelens JJ. Infection with a respiratory virus before hematopoietic cell transplantation is associated with alloimmune-mediated lung syndromes. *J Allergy Clin Immunol* 2018;141:697–703.

E2.	Zinter MS, Lindemans CA, Versluys BA, Mayday MY, Sunshine S, Reyes G, Sirota M, Sapru A, Matthay MA, Kharbanda S, Dvorak CC, Boelens JJ, DeRisi JL. The pulmonary metatranscriptome prior to pediatric HCT identifies post-HCT lung injury. *Blood* 2021;137:1679–1689.

E3.	Versluys AB, van der Ent K, Boelens JJ, Wolfs T, de Jong P, Bierings MB. High Diagnostic Yield of Dedicated Pulmonary Screening before Hematopoietic Cell Transplantation in Children. *Biol Blood Marrow Transplant* 2015;21:1622–1626.

E4.	Versluys AB, Rossen JW, van Ewijk B, Schuurman R, Bierings MB, Boelens JJ. Strong association between respiratory viral infection early after hematopoietic stem cell transplantation and the development of life-threatening acute and chronic alloimmune lung syndromes. *Biol Blood Marrow Transplant* 2010;16:782–791.

E5.	Panoskaltsis-Mortari A, Griese M, Madtes DK, Belperio JA, Haddad IY, Folz RJ, Cooke KR, American Thoracic Society Committee on Idiopathic Pneumonia Syndrome. An official American Thoracic Society research statement: noninfectious lung injury after hematopoietic stem cell transplantation: idiopathic pneumonia syndrome. *Am J Respir Crit Care Med* 2011;183:1262–1279.

E6.	Jagasia MH, Greinix HT, Arora M, Williams KM, Wolff D, Cowen EW, Palmer J, Weisdorf D, Treister NS, Cheng GS, Kerr H, Stratton P, Duarte RF, McDonald GB, Inamoto Y, Vigorito A, Arai S, Datiles MB, Jacobsohn D, Heller T, Kitko CL, Mitchell SA, Martin PJ, Shulman H, Wu RS, Cutler CS, Vogelsang GB, Lee SJ, Pavletic SZ, *et al.* National Institutes of Health Consensus Development Project on Criteria for Clinical Trials in Chronic Graft-versus-Host Disease: I. The 2014 Diagnosis and Staging Working Group report. *Biol Blood Marrow Transplant* 2015;21:389–401.

E7.	Stanojevic S, Graham BL, Cooper BG, Thompson BR, Carter KW, Francis RW, Hall GL, Global Lung Function Initiative TLCO working group, Global Lung Function Initiative (GLI) TLCO. Official ERS technical standards: Global Lung Function Initiative reference values for the carbon monoxide transfer factor for Caucasians. *Eur Respir J* 2017;50:.

E8.	Culver BH, Graham BL, Coates AL, Wanger J, Berry CE, Clarke PK, Hallstrand TS, Hankinson JL, Kaminsky DA, MacIntyre NR, McCormack MC, Rosenfeld M, Stanojevic S, Weiner DJ, ATS Committee on Proficiency Standards for Pulmonary Function Laboratories. Recommendations for a Standardized Pulmonary Function Report. An Official American Thoracic Society Technical Statement. *Am J Respir Crit Care Med* 2017;196:1463–1472.

E9.	Koopman M, Zanen P, Kruitwagen CLJJ, van der Ent CK, Arets HGM. Reference values for paediatric pulmonary function testing: The Utrecht dataset. *Respir Med* 2011;105:15–23.

E10.	Quanjer PH, Stanojevic S, Cole TJ, Baur X, Hall GL, Culver BH, Enright PL, Hankinson JL, Ip MSM, Zheng J, Stocks J, ERS Global Lung Function Initiative. Multi-ethnic reference values for spirometry for the 3-95-yr age range: the global lung function 2012 equations. *Eur Respir J* 2012;40:1324–1343.

E11.	Zinter MS, Dvorak CC, Mayday MY, Iwanaga K, Ly NP, McGarry ME, Church GD, Faricy LE, Rowan CM, Hume JR, Steiner ME, Crawford ED, Langelier C, Kalantar K, Chow ED, Miller S, Shimano K, Melton A, Yanik GA, Sapru A, DeRisi JL. Pulmonary Metagenomic Sequencing Suggests Missed Infections in Immunocompromised Children. *Clin Infect Dis* 2019;68:1847–1855.

E12.	Mayday MY, Khan LM, Chow ED, Zinter MS, DeRisi JL. Miniaturization and optimization of 384-well compatible RNA sequencing library preparation. *PLoS One* 2019;14:e0206194.

E13.	Kalantar KL, Carvalho T, de Bourcy CFA, Dimitrov B, Dingle G, Egger R, Han J, Holmes OB, Juan Y-F, King R, Kislyuk A, Lin MF, Mariano M, Morse T, Reynoso LV, Cruz DR, Sheu J, Tang J, Wang J, Zhang MA, Zhong E, Ahyong V, Lay S, Chea S, Bohl JA, Manning JE, Tato CM, DeRisi JL. IDseq-An open source cloud-based pipeline and analysis service for metagenomic pathogen detection and monitoring. *Gigascience* 2020;9:giaa111.

E14.	Davis NM, Proctor DM, Holmes SP, Relman DA, Callahan BJ. Simple statistical identification and removal of contaminant sequences in marker-gene and metagenomics data. *Microbiome* 2018;6:226–2.

E15.	Zinter MS, Mayday MY, Ryckman KK, Jelliffe-Pawlowski LL, DeRisi JL. Towards precision quantification of contamination in metagenomic sequencing experiments. *Microbiome* 2019;7:62–6.

E16.	Newman AM, Steen CB, Liu CL, Gentles AJ, Chaudhuri AA, Scherer F, Khodadoust MS, Esfahani MS, Luca BA, Steiner D, Diehn M, Alizadeh AA. Determining cell type abundance and expression from bulk tissues with digital cytometry. *Nat Biotechnol* 2019;37:773–782.
